# Dynamic interplay of polygenic risk across brain disorders, neuropathological endophenotypes, and neuropsychiatric symptoms

**DOI:** 10.64898/2026.06.30.26356924

**Authors:** N.J. Mekkes, S. Kumar, E. Hoekstra, A. Marmolejo-Garza, E. Dagkesamanskaia, D. Wever, M. Groot, K.L. Kreft, A. Rajicic, H. Seelaar, L. Donker Kaat, J. van Swieten, S.M.M. Vermorgen, A.J. Rozemuller, N. Fransen, H.J. Westra, B.J.L. Eggen, I. Huitinga, I.R. Holtman

## Abstract

Neurodegenerative and psychiatric brain disorders show substantial heterogeneity in clinical and neuropathological manifestations, possibly reflecting both distinct and shared pathogenic mechanisms. Our understanding of brain disorder heterogeneity is limited by the scarcity of deeply phenotyped, multi-modal post-mortem brain bank cohorts. Here, we integrated genotype, clinical and neuropathological data from 2,553 Netherlands Brain Bank donors to investigate how genetic risk contributes to disease manifestations. Disease-specific polygenic risk scores revealed extensive cross-disorder enrichment of genetic risk, suggesting shared pathogenic mechanisms beyond diagnostic boundaries. Donors with frontotemporal lobar degeneration carrying *C9orf72* repeat expansions showed elevated polygenic risk for multiple disorders, indicating that genetic variation may modify monogenic disease expression. Neuropsychiatric symptoms were associated with distinct personality-trait polygenic risk profiles across disorders, highlighting diagnosis-dependent genetic contributions to clinical heterogeneity. Together, our findings demonstrate the value of this unique resource and reveal a dynamic interplay of genetic risk across brain disorders.

## Introduction

Brain disorders, such as Alzheimer’s disease (AD), frontotemporal dementia (FTD), multiple sclerosis (MS), Parkinson’s disease (PD), and psychiatric illnesses, are highly multifactorial conditions whose underlying disease mechanisms remain incompletely understood. It remains challenging to predict who will develop a brain disorder^1,2^, to establish an accurate clinical diagnosis that corresponds to neuropathology^3–6^, and to make reliable prognoses about disease progression^7,8^. These challenges largely stem from the extensive clinical and neuropathological heterogeneity that characterizes brain disorders^9^. A single disorder can present with widely divergent clinical trajectories, sometimes mimicking other disease entities, and conversely, individuals with distinct diagnoses may share overlapping symptomatology^10^.

Neuropathological studies have similarly revealed substantial variation within and across disorders. While AD is classically defined by extracellular amyloid-β plaques and intracellular neurofibrillary tangles (NFT) of hyperphosphorylated tau, many individuals with AD pathology also harbor additional neuropathological endophenotypes^11,12^. Conversely, clinically unaffected individuals may present intermediate levels of these same pathologies^13^. Therefore, neurodegenerative disorders are increasingly viewed as states along multiple spectra of partly independent neuropathological endophenotypes, rather than a discrete entity with a mutually exclusive profile^9,10,14,15^. The full range of these endophenotypes can only be systematically captured through neuropathological investigation, underscoring the critical importance of brain banks not only as repositories of tissue, but also as integrative data resources that enable standardized, multi-modal characterization of brain disorders^9^.

Genetic studies play a central role in advancing our understanding of the biological basis of brain disorders. Genome-wide association studies (GWAS) aim to identify common genetic variants associated with complex traits, such as disease status or quantitative neuropathological endophenotypes^16,17^. These studies have demonstrated that most neurodegenerative and psychiatric conditions are highly polygenic. Cross-disorder analyses have revealed substantial shared genetic architectures among psychiatric disorders, while neurological disorders were less correlated but do share a subset of risk loci^18–23^. An additional level of complexity is added by GWAS by proxy which relies on family-reported assessments of disorders that may include a considerable number of misdiagnosed individuals^24^. A promising direction has been the application of GWAS to neuropathological endophenotypes^25^, which reflects the underlying disease biology more closely. However, the number of standardized, large-scale, post-mortem datasets with genetic data limits such efforts, and the specificity of these scores have not yet been assessed.

Post-GWAS studies subsequently aimed to identify the molecular and cellular mechanisms by which GWAS risk loci exert their effects. Genetic risk for AD has been linked to microglia enhancers^26,27^, PD risk to lysosomal function^27^, and MS risk to adaptive immune-related pathways^28^. Together, these findings suggest that GWAS signals reflect underlying neurobiological mechanisms. However, it remains unclear to what extent these mechanisms are disorder-specific or represent shared processes across brain disorders.

Here, we present comprehensive genetic analyses of brain donors from the Netherlands Brain Bank (NBB) within the framework of the Netherlands Neurogenomics Database (NND), integrating clinical trajectories, neuropathological endophenotypes, and genetic data from 2,553 brain donors. We combined genetic data with neuropathological examinations and clinical trajectories to (i) perform polygenic risk score (PRS) comparisons across diagnostic entities and neuropathological endophenotypes, (ii) analysis of genetic interactions in familial frontotemporal lobar degeneration (FTLD), and motor neuron disease (MND) spectrum disorder mutation carriers, (iii) conduct GWAS on neuropathologically defined case-control status to refine existing GWAS, (iv) investigate associations between personality traits-PRS and neuropsychiatric symptoms across diagnosis groups. Together, these analyses demonstrate how a deeply phenotyped brain bank can refine and contextualize signals from large-scale GWAS and give new insight into disease mechanisms.

## Results

### Population structure of the cohort

To better understand the genetic structure associated with neuropathology, we performed genotyping on 2,802 donors (Extended Data Fig. 1a) and processed 2,667 neuropathological reports (Extended Data Fig. 1b). For a graphical overview of the workflow and analyses, see Fig. 1a. After applying quality control and filtering procedures, we retained 2,553 donors with high-quality genetic data across 34 neuropathologically defined disorder groups (Fig. 1b, Extended Data Table 1, Supplementary Table 1). The cohort originates from all around the Netherlands (Fig. 1c). To assess population structure, we performed clustering on population stratification results (Fig. 1c), and observed subtle differences related to regional variation. These results confirm that we have obtained a high-quality genetic dataset and that the cohort reflects regional variation in the Dutch population^29^.

**Fig. 1.**
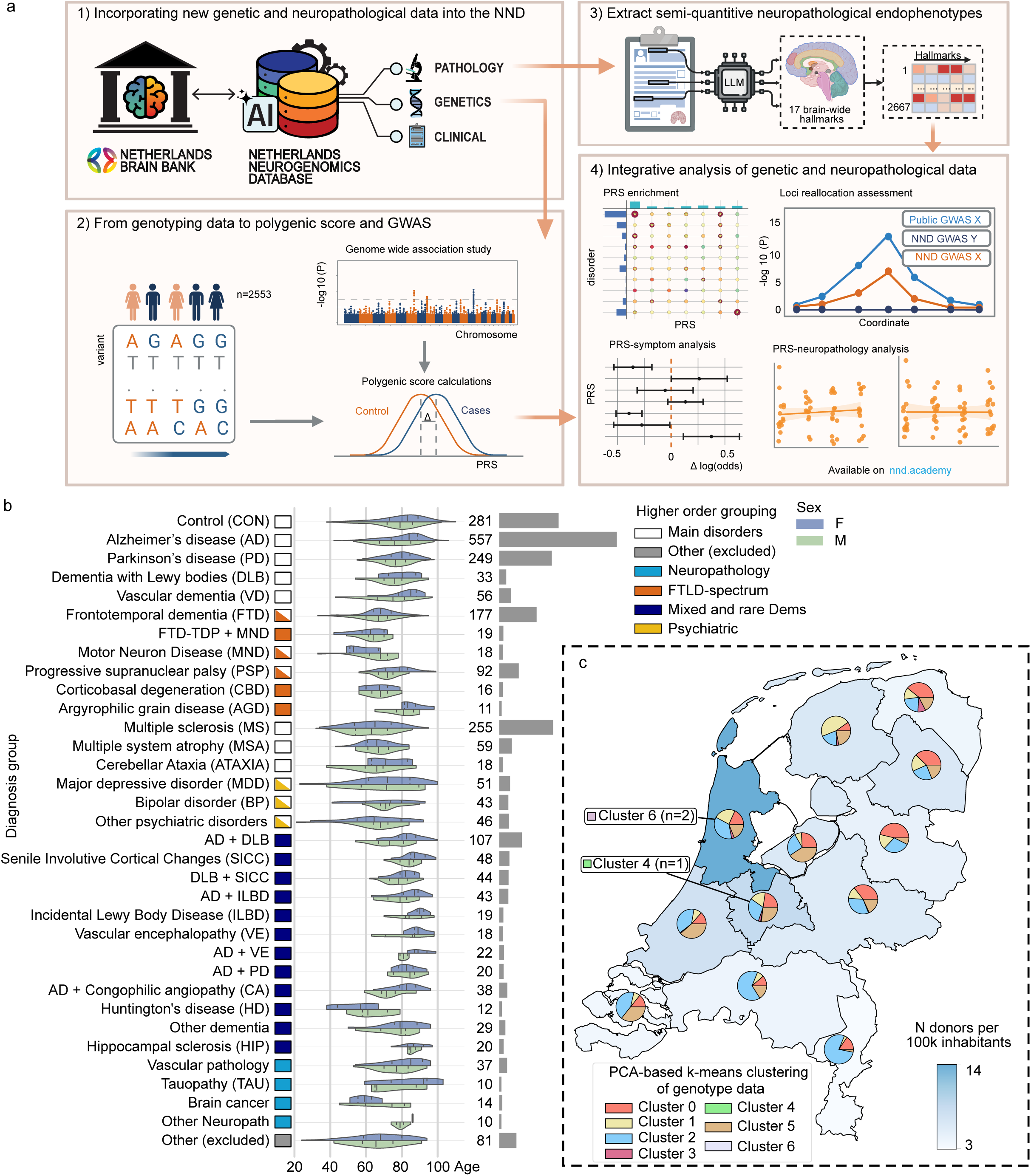
a, Overview of the study workflow. Starting from the NND (a1), genotype data was collected and processed (a2), as were neuropathological endophenotypes (a3). The main analytical approaches are summarized in a4, including PRS enrichments, symptom–PRS and neuropathology-PRS association testing, and locus refinement. b, Cohort composition shown as violin plot split by sex for each diagnosis, with sample sizes with proportional bars on the right. Boxes indicate higher order groupings for analysis purposes. c, Geographic distribution of donors by province (heatmap indicating n donors per 100.000 inhabitants), with pie charts showing proportions of k-means clusters per province based on genotype-derived principal components.

### Many PRS for brain disorders are pleiotropic, suggesting shared genetic mechanisms

To investigate the genetic pleiotropy of brain disorders, we collected previously published GWAS summary statistics for 11 neurodegenerative and psychiatric disorders^30–40^ (Extended Data Table 2). We subsequently calculated and compared PRS distributions using SBayesRC^41^ between cases and controls across 13 most common diagnostic groups (Fig. 2a-d, Supplementary Table 2). As expected, the AD group exhibited a significantly higher AD-PRS compared to controls (Fig. 2a). Similar patterns were observed for FTD, and MS with increased FTD-PRS and MS-PRS compared to controls, respectively (Fig. 2b,c).

**Fig. 2.**
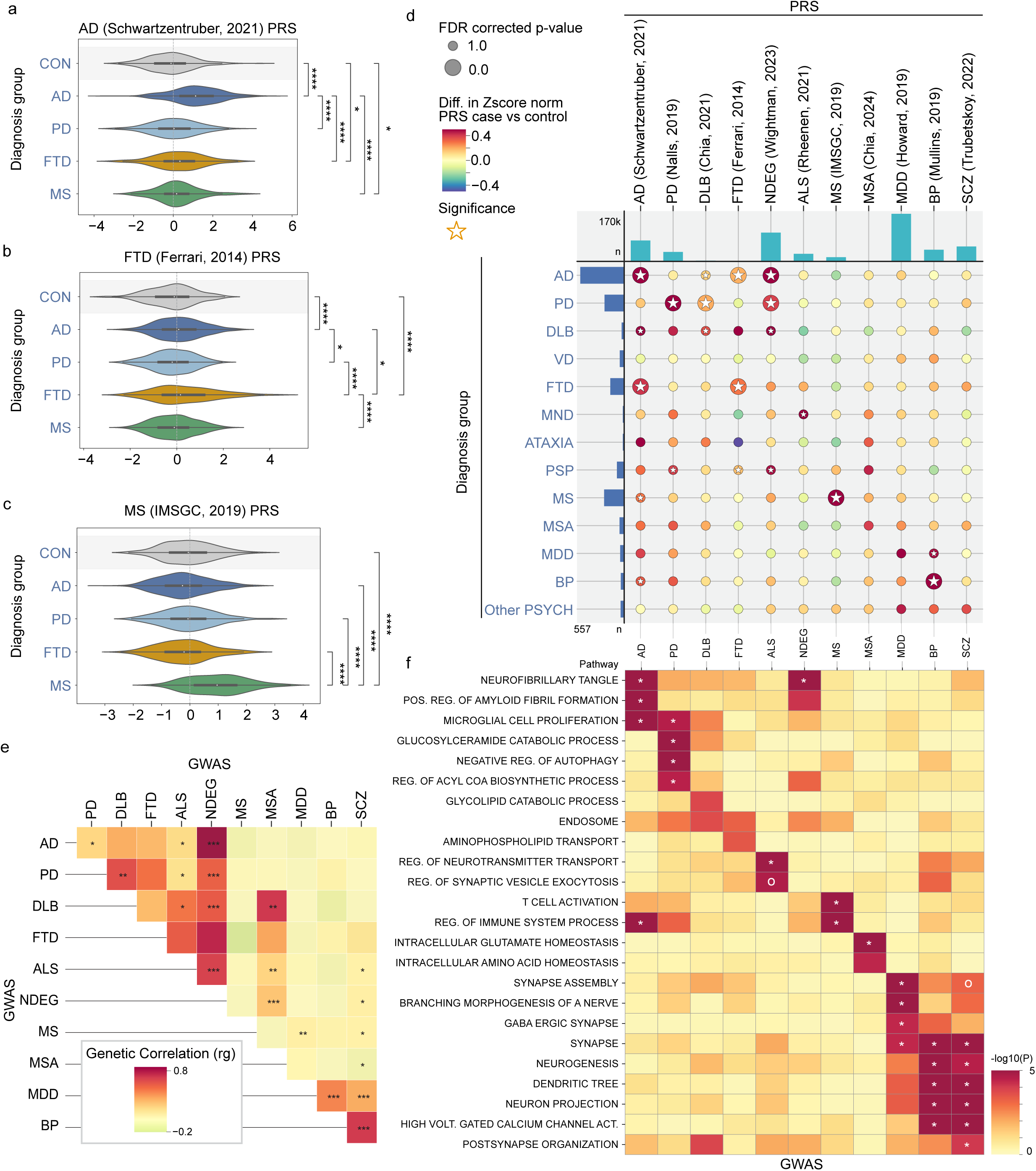
a-c, Violin plots show the distribution of AD-PRS, FTD-PRS, and MS-PRS in donors grouped by neuropathological diagnosis (CON, AD, PD, FTD, and MS). Pairwise statistical comparisons were performed between all groups (FDR corrected permutation test, *P < 0.05, **P < 0.005, ***P<0.0005, ****P < 0.00005). d, Integrated dot plot depicting the delta-PRS for 11 disorders (columns) for 13 neuropathological diagnosis groups (rows). Color indicates the direction and magnitude of the difference in standardized PRS (case vs control). Dot size reflects statistical significance, and stars denote FDR-corrected permutation test P < 0.05. Column bar plot indicates GWAS sample size (n cases). Row bar plots indicate group sample size. e, Heatmap of pairwise genetic correlations between collected GWAS sumstats, estimated using LDSC, with stars denoting FDR corrected significance levels. f, Heatmap depicting the MAGMA gene-set enrichment results for each PRS, showing –10 log FDR-corrected P-values for selected gene ontology sets (*p<0.05, °p<0.1).

Next, we compared PRS across multiple groups (Fig. 2d, Extended Data Fig. 2a) and found that most GWASs are enriched in the corresponding disorder. For example, PD-PRS is enriched in PD, Amyotrophic lateral sclerosis (ALS)-PRS in the MND group (diagnostic group including ALS and other MND donors), BP-PRS in the BP-group. However, this comparison could not be performed for Vascular Dementia (VD), cerebellar ataxia, or progressive supranuclear palsy (PSP) donors because disorder-specific PRS were not available. Multiple system atrophy (MSA) showed a higher but non-significant MSA-PRS relative to controls, possibly due to the lower sample size of this GWAS. AD, PD, MND, MS, Major depressive disorder (MDD), and BP also had the highest median values for their corresponding disorder PRS (Extended Data Fig. 2a).

Interestingly, the AD-PRS was also significantly higher in Dementia with Lewy bodies (DLB), FTD, MS, and Bipolar disorder (BP) donors when compared to controls, suggesting that genetic features underlying AD susceptibility might also contribute to the risk of other disorders in a subset of donors. Genetic correlation analysis of the GWAS summary statistics using Linkage Disequilibrium Score Regression (LDSC^42, 43^) showed no significant global association between AD and DLB, FTD, MS, or BP (Fig. 2e). In line with previous findings^30^, the AD summary stats were enriched for pathways related to NFT, amyloid fibril formation, and microglial cell proliferation (Fig. 2f, Supplementary Table 3). The enrichment of the AD-PRS in other disorders suggests that these biological processes might also contribute to other disorders, outside of the scope in which they were originally identified. For example, novel lines of evidence suggest that tau and microglia cell proliferation might also contribute to MS progression and susceptibility^44–46^; our genetic results might offer further evidence to substantiate this. It also shows the promise of a cross-disorder approach to study heterogeneity across related disorders to obtain a new insight into potentially relevant disease mechanisms.

In addition, we calculated a general neurodegeneration-PRS (NDEG-PRS) based on previously published genomic structural equation model^47^ summary statistics capturing genetic liability shared across AD, ALS, DLB, and PD^34^. This NDEG-PRS is enriched in AD, PD, DLB, and PSP groups, but surprisingly not significantly in MND, and neither in VD, FTD or MSA suggesting that this polygenic signal might not generalize to all forms of neurodegeneration.

As many patients with neurodegenerative disorders do not fall into neatly defined neuropathological categories due to comorbidity, we investigated whether PRS could help characterize atypical and mixed forms of dementia (Extended Data Fig. 2b, Supplementary Table 4). Of the 12 rare and mixed dementia groups, only six showed a significantly higher AD-PRS compared to controls. Three of these also showed an enrichment for PD, DLB, and or FTD-PRS, corroborating the dynamic interplay between polygenic risk across brain disorders.

Another noteworthy observation was that the MDD donors exhibited significantly higher BP-PRS, but not MDD-PRS. When stratifying the analysis by sex (Extended Data Fig. 2c,d, Supplementary Table 5 and Supplementary Table 6), we observed a clear enrichment of MDD-PRS in male MDD donors, which was absent in female MMD donors. This suggests that the male donors more closely reflect the MDD-specific genetic profiles than the females. Consistent with this interpretation, previous studies report sex differences in the genetic architecture of MDD^48^, with evidence for higher heritability in females. Sex specific stratification analysis of disease PRS also showed that FTD-PRS is significantly enriched in both AD and FTD females, but not in males, suggesting that this polygenic signal may play a more prominent role in females than in males.

### Polygenic risk across FTLD–MND spectrum subtypes

The FTLD-MND spectrum comprises a diverse group of neurodegenerative disorders that predominantly affect the frontal and temporal lobes and/or spinal cord, often with aggregation of TDP-43 or tau. Neuropathological subclassifications are made based on the morphological characteristics of the inclusions, and include among others different TDP subtypes (TDP-A, TDP-B and TDP-C), Pick’s disease (PID), PSP, Argyrophilic grain disease (AGD), and corticobasal degeneration (CBD). It is currently not well understood which genetic factors contribute to susceptibility to these different subtypes. To address this question, we made use of the same set of PRSs for brain disorders, and compared the distributions of each group to controls.

Out of 12 FTLD-MND spectrum disorders, six exhibited a significant enrichment for FTD-PRS, including FTD-TDP-B, FTD-TDP-C, PSP, CBD, and FTD-PID (Fig. 3a, Supplementary Table 7), while three FTLD disorders exhibited enrichments for other PRSs. Since FTD as a parent category had a significantly higher AD PRS (Fig. 2d), we first assessed which FTLD subtypes were enriched for AD-PRS. Two subtypes (FTD-TDP-A, FTD-TDP-B) showed significant AD-PRS enrichment, suggesting that the genetic features of AD, such as microglia proliferation, might also contribute to susceptibility of these FTD subtypes. The PD-PRS was enriched in the PSP subtype only, suggesting the PD-PRS signal might also reflect genetic risk related to nigra-striatal degeneration, a shared feature of these two disorders. These findings could place the clinical overlap between PSP and PD in a new light.

**Fig. 3.**
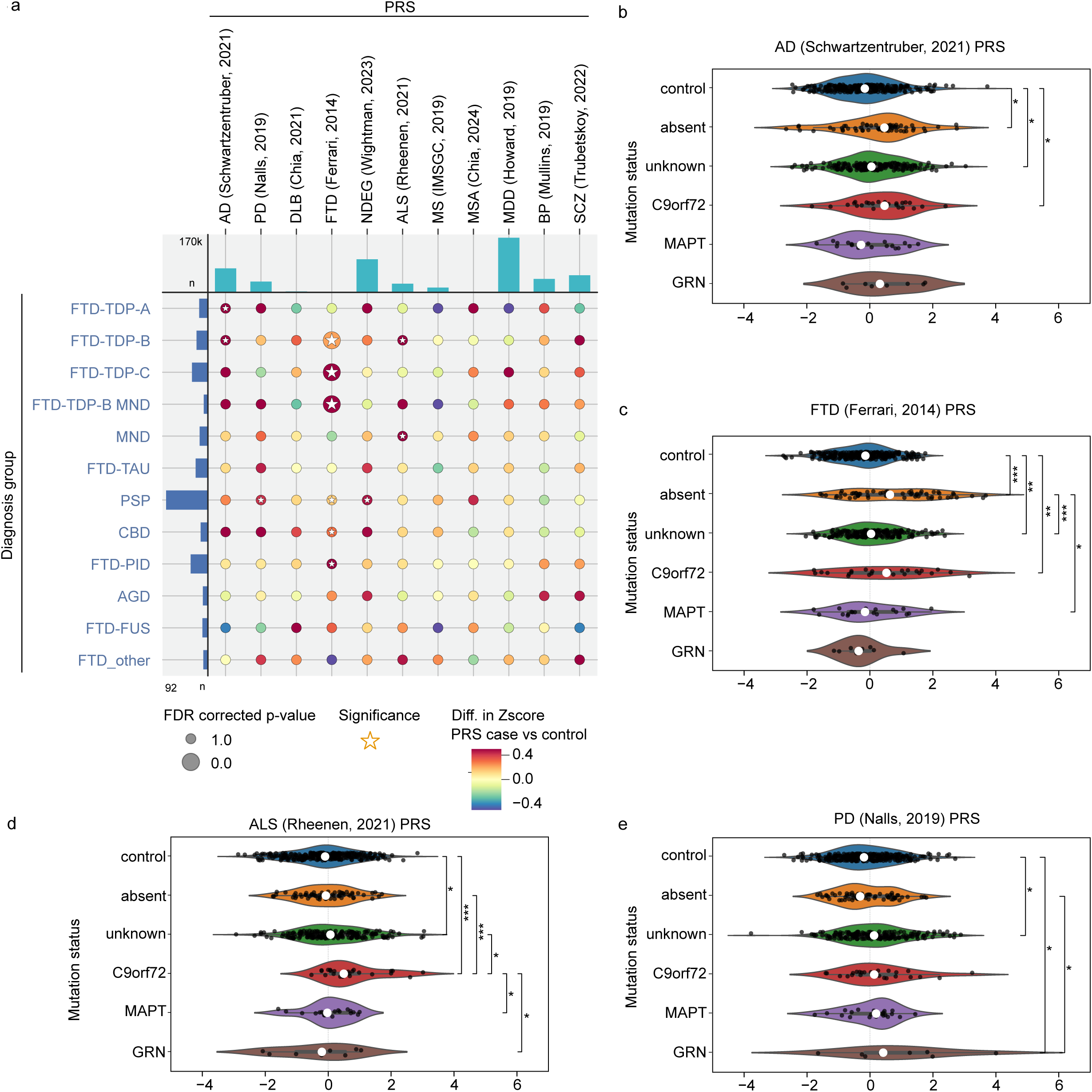
a, Integrated dot-plot depicting delta PRS for 11 different disorders (columns) for 12 FTLD-MND spectrum subtypes (rows). Color indicates the direction and magnitude of the difference in standardized PRS (case vs control). Dot size reflects statistical significance, and stars denote FDR-corrected permutation test P < 0.05. Column bar plot indicates GWAS sample size (n cases). Row bar plots indicate group sample size. b-e, Violin plots show the distribution of AD, FTD, ALS, and PD PRS in donors grouped by mutation status (control donors, FTLD-MND spectrum donors with mutations absent, unknown, in C9orf72, in MAPT, in GRN). Pairwise statistical comparisons were performed between each group (FDR corrected permutation test, *P < 0.05, **P < 0.005, ***P < 0.0005).

### FTLD-MND spectrum disorders familial mutations and PRS interaction

FTLD-MND spectrum disorders are frequently associated with familial mutations, including those in the *MAPT*, *GRN*, and *C9orf72* genes^49^. Increasing evidence from other disorders suggest that polygenic risk and familial mutations can interact and influence disease phenotypes^50, 51^. Whether this also applies to FTLD-MND spectrum related mutations was unknown. To test polygenic–monogenic interactions, we compared FTD-PRS, ALS-PRS, PD-PRS, and AD PRS within the FTLD-MND spectrum group, taking mutation status into account (Fig 3b-e, Supplementary Table 8). Mutation status was known for 181 of our 373 FTLD-MND donors^52^. Here we observed that donors with a hexarepeat expansion in the *C9orf72* gene, which is often linked to FTD-TDP-B, had a significantly elevated PRS for FTD, AD, and ALS but not for PD when compared to controls. Moreover, donors with a *GRN* pathogenic variant showed an increased PD PRS, but not FTD, AD, or ALS PRS. This finding supports a dynamic interplay between disease-associated mutations and polygenic risk.

### GWAS refinement through GWAS on neuropathologically diagnosed cohort

Many large-scale GWASs for brain disorders rely on clinical instead of pathologically defined diagnoses and familial proxy cases, and as a result, some loci may be driven by clinically misdiagnosed individuals. We hypothesized that our neuropathologically defined cohort could help evaluate the disease specificity of these associations and identify loci that may be better attributed to other disorders. We therefore performed case-control GWAS for AD, FTD, and PD (Extended Data Fig. 3a-f) and systematically interrogated 37 genome-wide significant AD loci reported by^30^. For each locus, we conducted colocalization analysis^53^ between the published AD GWAS and our neuropathological AD, FTD, and PD GWASs to assess disease specificity (Coloc PP4, Fig. 4a). Most AD_Schwartzentruber_ GWAS risk loci colocalized most strongly with neuropathological AD_NND_ GWAS. However, a locus on chromosome 7 near *EPHA1* showed strong evidence of colocalization with FTD (PP4 = 0.952, PP3 = 0.001) but only weak evidence with AD (PP4 = 0.390, PP3 = 0.002) (Fig 4a,b, Supplementary Table 9). Although not genome-wide significant, the lead FTD variant (rs7810606) remained significant after locus-level multiple-testing correction (Supplementary Table 10). These findings suggest that the *EPHA1* locus may also contribute to FTD risk and that its reported association with AD could partly reflect misdiagnosed FTD cases in clinically defined cohorts.

**Fig. 4.**
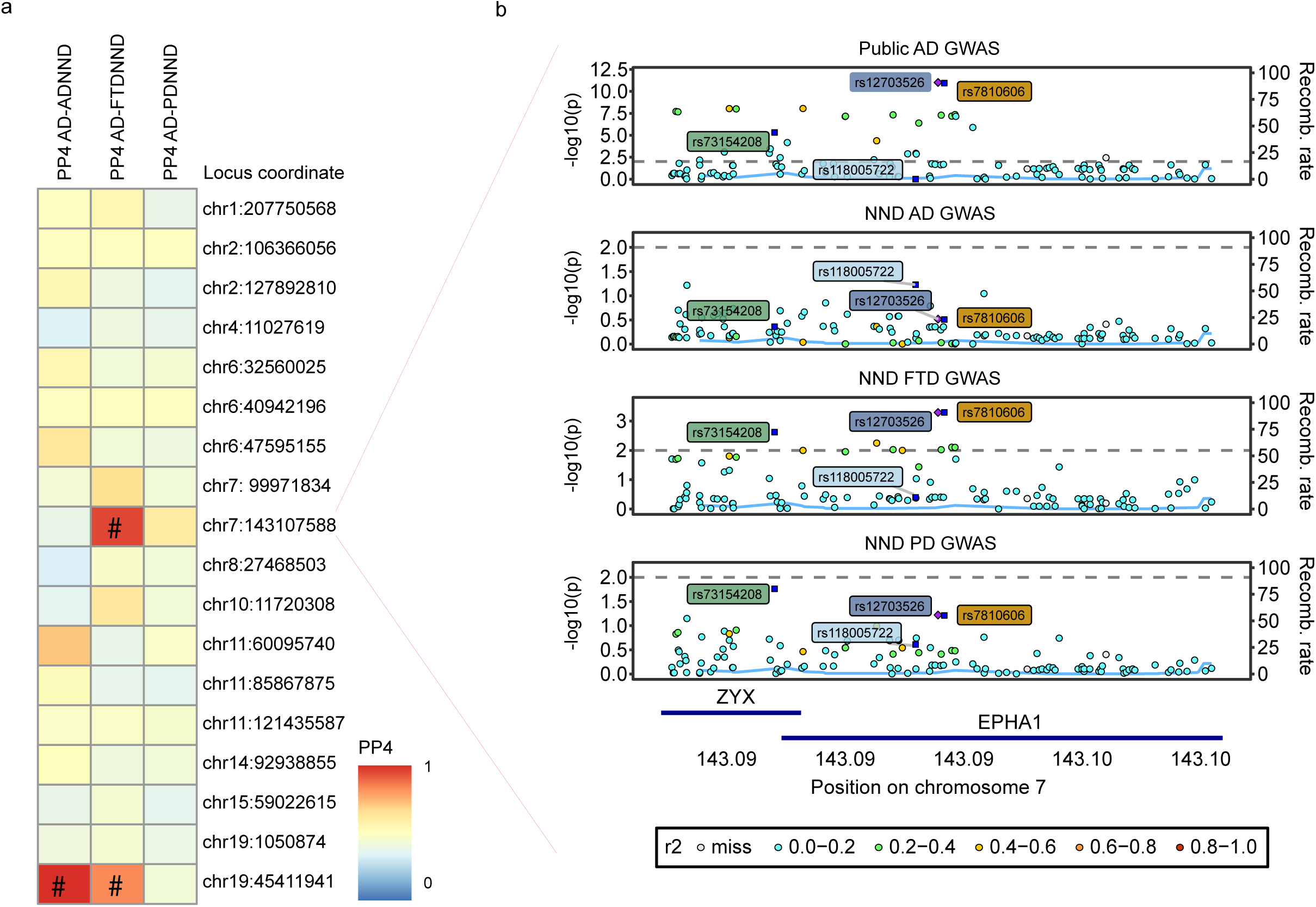
a, Heatmap showing AD risk loci colocalization results of previously published summary statistics (Schwartzentruber et al. (2022)) and summary statistics from Alzheimer’s disease (AD), frontotemporal dementia (FTD), and Parkinson’s disease (PD) in the NBB cohort. Rows represent AD loci de ned in Schwartzentruber et al. (2022), columns indicate pairwise comparisons between each pathological GWAS and the reference AD GWAS. Color denotes the posterior probability of a shared causal variant (PP4), with higher values indicating stronger evidence for colocalization (# PP4 > 0.80). b, Regional association plot for the chromosome 7 EPHA1 locus. The four tracks show association results from Schwartzentruber AD (lead SNP dark blue), NND AD (lead SNP lightblue), NND FTD (lead SNP orange), and NND PD GWAS (lead SNP green), respectively. The left y axis shows −log (P), and the right y axis the recombination rate. Variants are colored by LD (r²) with the lead SNP, with gene annotations below. The locus is genome-wide signi cant in the reference AD GWAS, absent in pathological AD, and shows a sub–genome-wide but signi cant signal in pathological FTD.

### Associations between neuropathological endophenotype score and disorder PRS

To characterize the neuropathological heterogeneity across brain disorders, we used a human-in-the-loop workflow supporting the Llama 3.1 8B Instruct large language model (LLM)^54^ to extract 17 neuropathological endophenotypes for 2,667 donors (Extended Data Fig. 1b, Extended Data Table 3). The most frequently observed neuropathological endophenotypes were atherosclerosis, infarctions, and hippocampus and amygdala atrophy, suggesting that vascular and structural changes are common contributors to the neuropathological burden in this cohort (Extended Data Fig. 4a). Pairwise comparison of neuropathological endophenotypes showed strong positive correlations among Alzheimer-type features, including amyloid, tau, neuritic plaque, and CAA-related measures. In contrast, vascular, α-synuclein, and structural abnormalities showed weaker and more variable associations, highlighting the heterogeneous composition of pathological burden across donors (Extended Data Fig. 4a). As expected, across diagnostic groups, neuropathological burden showed disease-specific patterns relative to controls. AD had higher Alzheimer-type pathology, including amyloid spread, tau/NFT burden, neuritic plaques, cerebral amyloid angiopathy (CAA) presence, and amygdala and hippocampus atrophy. AD also showed high presence of atherosclerosis, supporting the growing recognition that vascular pathology contributes to AD alongside amyloid-and tau-related processes^55–57^. PD and DLB were distinguished by higher α-synuclein/Lewy body burden and also showed modest increases in neurofibrillary tangle pathology, consistent with the frequent co-occurrence of Alzheimer-type changes in synucleinopathies, while VD showed higher atherosclerosis and ischemic injuries; hippocampus atrophy was also more common in FTD. MS was associated with atherosclerosis and corpus callosum atrophy (Extended Data Fig. 4b, Supplementary Table 11). Together these observations are consistent with expected neuropathological patterns, while also uncovering novel relationships, thereby supporting the high quality of our pathology dataset.

Next, we evaluated the associations between disease-specific PRS and neuropathology endophenotypes to understand how genetic risk relates to brain pathology. Overall, the patterns observed were biologically consistent and showed that genetic risk is most strongly linked to core AD-related pathologies (Fig 5a-d, Supplementary Table 12). Overall, the AD PRS showed the strongest and most consistent associations with multiple neuropathological endophenotypes. Higher AD genetic risk was positively and significantly associated with increased amyloid burden and stage, NFT burden and stage, CAA and CAA-severity, and hippocampus atrophy. Interestingly, even though atherosclerosis was strongly increased in AD donors, it was not significantly associated with the AD-PRS. This suggests that the vascular component of the AD pathology might be more strongly influenced by environmental factors, than by common genetic risk for AD. These findings align with known AD biology and indicate that genetic risk for AD is reflected in its hallmark pathological endophenotypes. Associations with Alpha-synuclein spread showed non-significant trends for PD PRS, and DLB PRS, suggesting that genetic risk for these conditions may not translate directly into α-synuclein spread or burden.

**Fig. 5.**
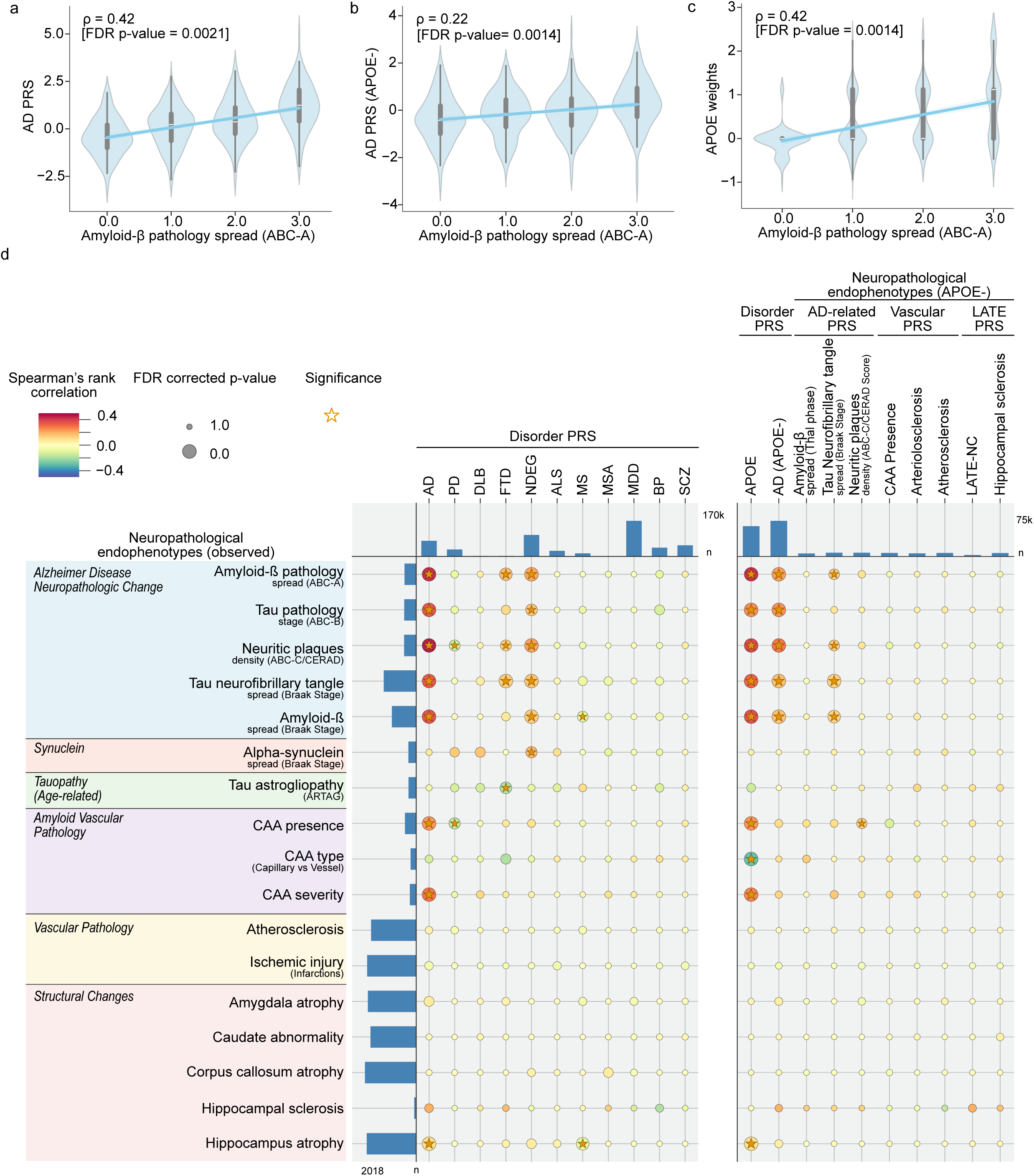
Associations between polygenic (risk) scores and neuropathological endophenotype. a-c, Scatter plots with trend lines showing Spearman rank correlation (ρ) between standardized polygenic (risk) score and neuropathological endophenotype. Statistical significance was assessed using permutation testing with FDR-correction, depicted as P values. d, Integrated dot plot summarizing associations between PRS for 11 neurological and neurodegenerative disorders (columns, left), APOE-related scores including APOE and AD-PRS excluding the APOE region, and 8 neuropathological endophenotypes (columns, right) and 17 neuropathological endophenotypes (rows). Dot color represents the direction and magnitude of the correlation, with red indicating positive and blue indicating negative associations. Dot size reflects statistical significance, and stars denote associations passing FDR-corrected permutation significance (P<0.05). The top bar plot shows GWAS cohort sizes (number of cases) for each PRS, while the side bar plot indicates sample size availability for each neuropathological endophenotype.

A recent GWAS of multiple neuropathological endophenotypes provided an opportunity to further dissect the genetic factors underlying neuropathological variation^25^. Based on this study, we calculated neuropathology endophenotype PRS and aimed to assess if these PRSs were associated with their corresponding neuropathological endophenotypes as a means to evaluate their specificity (Fig. 5d, Supplementary Table 13). APOE showed the strongest and broadest associations across AD-related neuropathological endophenotypes, including CAA, and hippocampus atrophy. AD-PRS showed a more specific pattern aligned with AD neuropathological change. In contrast, *APOE*-excluded endophenotype PRS showed weaker signals, with limited evidence for endophenotype-specific effects. These findings suggest that *APOE* captures much of the shared AD pathological burden, whereas current endophenotype PRS have limited ability to disentangle highly correlated neuropathological endophenotypes in endstage disease.

### Polygenic trait scores contribute to symptom-level heterogeneity

Patients with neurodegenerative disorders show marked heterogeneity in neuropsychiatric symptoms, but the biological basis of this variability remains poorly understood^58^. We tested whether polygenic scores for neuropsychiatric disorders and personality traits (risk taking^59^, intelligence^60^, insomnia^59^, sociability^61^, and neuroticism^62^) contribute to symptom-level heterogeneity within neuropathologically defined diagnoses. For four disorder groups, we modeled the association between nine polygenic scores and the presence versus absence of nine specific psychiatric symptoms (Fig. 6, Supplementary Table 14, 15).

**Fig. 6.**
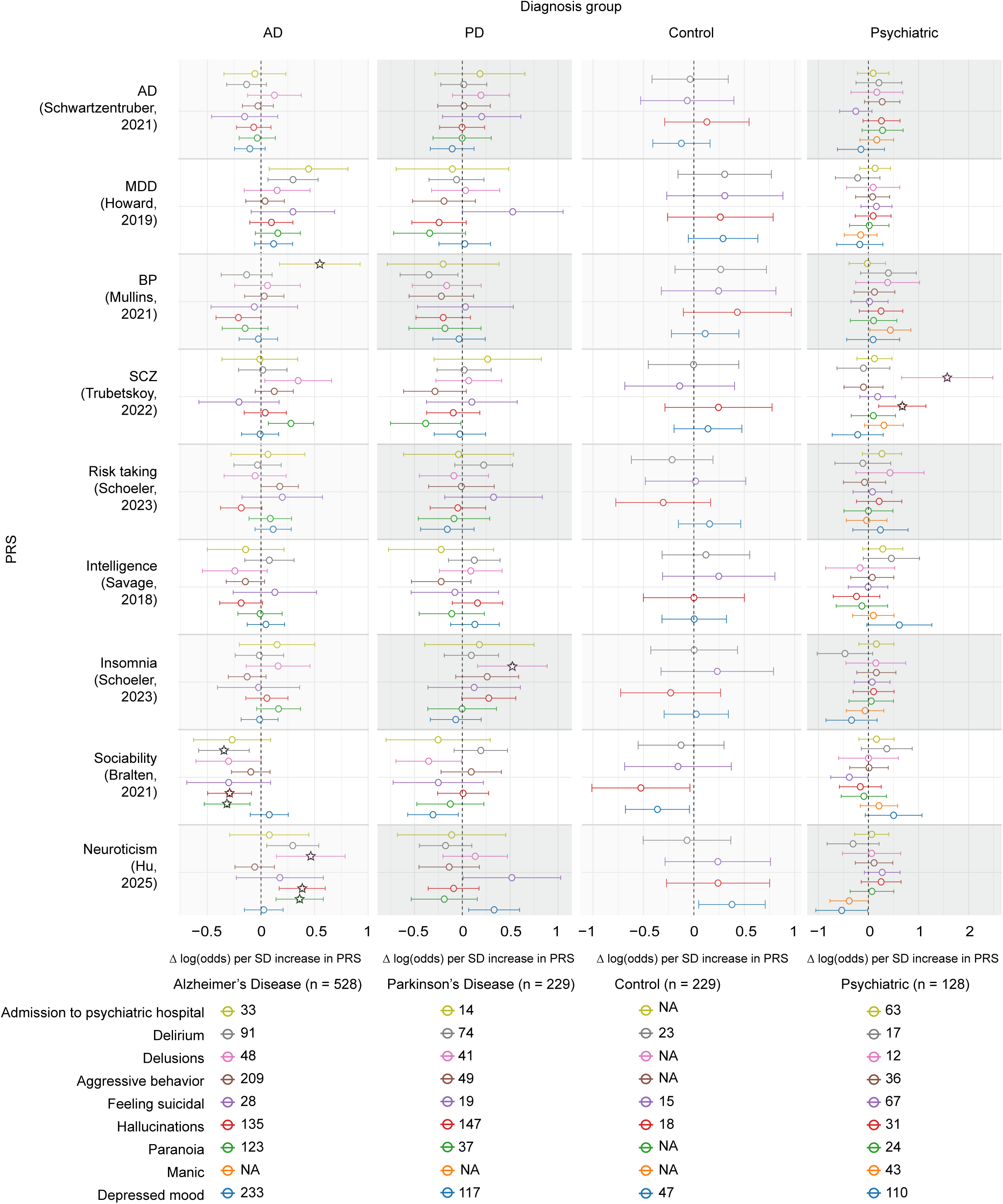
Forest plots for AD, PD, controls, and psychiatric donors showing the change in log odds per standard deviation increase in polygenic (risk) score (x-axis) for Alzheimer’s disease, major depressive disorder, bipolar disorder, schizophrenia, risk-taking, intelligence, insomnia, sociability, and neuroticism, across nine psychiatric domain symptoms (admission to psychiatric hospital, delirium, delusions, aggressive behavior, feeling suicidal, hallucinations, paranoia, manic, depressed mood). Sample size is indicated below each disorder column. Significant symptom-PRS interactions (passing permutation test threshold) are indicated with a star.

Among donors with AD, hallucinations, delusions, and paranoia were significantly associated with higher neuroticism polygenic scores, while hallucinations and delirium are significantly associated with lower sociability polygenic scores, suggesting that personality traits contribute to one’s likelihood of developing neuropsychiatric symptoms in AD. This is in line with previous publications^58, 63, 64^. These findings were not replicated in the PD-group, where, on the contrary, delusions were significantly associated with insomnia-PRS. Among psychiatric donors, hallucinations or delusions were significantly associated with an increased schizophrenia-PRS (SCZ-PRS). This is particularly interesting given the high genetic correlation between SCZ and BP, and the observation that the SCZ donors did not exhibit a significantly higher SCZ PRS compared to control (data not shown). Taken together, this suggests a link between neuropsychiatric symptoms and neuroticism and sociability in AD, while in PD the insomnia PRS plays a role, and among psychiatric donors the same symptoms are linked with high SCZ-PRS.

## Discussion

Here we present the Netherlands Neurogenomics database, a unique multi-modal data resource that includes genetic, neuropathological, and clinical data of post-mortem brain donors from the Netherlands Brain Bank. We calculated PRS for brain disorders, neuropathological endophenotypes, and personality traits, and compared them within and across neuropathologically defined disease groups. We were able to link genetic susceptibility directly to disease diagnoses and the accompanying clinical phenotypes. This approach circumvents one of the major limitations of current large-scale GWAS that rely on clinical diagnoses and proxy cases, where diagnostic heterogeneity and misclassification can obscure genuine biological relationships^65^.

There is extensive literature using genetic correlations between GWAS datasets to examine unique and shared genetic risk across brain disorders^17–23^. Here, we used a complementary approach by directly calculating disease PRS at the donor level and comparing those across diagnostic groups. Our analysis partially recapitulated the genetic correlations, but also revealed new associations. For example, we found an increased AD-PRS in several other disorders including DLB, FTD, MS, and BP. While our genetic correlations analysis only showed a significant relation between AD and PD, AD and ALS. These differences may reflect the distinct nature of the two approaches, and might capture complementary aspects of the shared genetic architecture, possibly due to different diagnostic inclusion criteria.

Nonetheless, some interesting observations are worthwhile exploring in more detail. For instance, donors with MS exhibited a small but significant increase in AD-PRS, suggesting that susceptibility to AD might partially capture biological processes relevant beyond AD itself. The AD GWAS score is particularly enriched for microglia, NFT, and amyloid plaques, raising the possibility that AD associated microglia subsets and or tau related pathways may contribute to MS pathophysiology, in a subset of individuals. This interpretation is consistent with neuropathological evidence implicating tau pathology might be involved in slow progression and cognitive decline in MS patients^66–69^. Interestingly, while the genome-wide genetic correlation between AD and MS is negligible, shared genetic loci were identified, offering additional evidence for a genetic link between these disorders^70^. The AD PRS score might explain part of the heterogeneity in MS neuropathology, why some donors show cognitive decline and slow progression of symptoms independent of relapse activity^71^.

Stratification of PRS across FTLD-MND spectrum subtypes suggested that polygenic risk differs by subtype and could potentially contribute to heterogeneity in FTLD subtype manifestation. Furthermore, the interaction between *C9orf72* hexarepeat expansion status and elevated FTD, AD and ALS PRS suggests that common variant burden can modulate the penetrance and expressivity of rare, high-impact mutations in the *C9orf72* gene but not in *MAPT*. *C9orf72* repeat expansions are among the most phenotypically heterogeneous genetic causes of FTLD, with substantial variability in clinical presentation and age at onset^72–74^. Our findings suggest that this heterogeneity might partially be shaped by the polygenic background. This aligns with recent views that for some disorders polygenic risk interacts with pathogenic mutations, with polygenic background shaping phenotypic outcomes in mutation carriers^75–77^.

Pathology-based GWAS and colocalization analyses demonstrated the value of brain bank cohorts for refining locus interpretation. At the *EPHA1* locus, we found stronger colocalization between a public AD GWAS and neuropathologically defined FTD than AD, suggesting either pleiotropic effects or misclassification of FTD cases in clinically defined AD cohorts. Research suggests that the *EPHA1* risk locus may impact the blood-brain barrier^78^ which could also play a role in FTD. As *EPHA1* has not previously been implicated in FTD, this finding highlights how neuropathologically verified cohorts can distinguish genuine pleiotropy from associations driven by diagnostic overlap.

To further disentangle whether cross-disorder genetic links reflect shared biology or mixed pathological burden, we examined neuropathological endophenotypes across diagnostic groups. These findings suggest that such genetic signals may partly arise from shared or co-occurring pathological processes, rather than diagnostic overlap alone. The modest NFT enrichment in PD and DLB supports the possibility that Alzheimer-type co-pathology contributes to heterogeneity within synucleinopathies. The distinct patterns of hippocampus and amygdala atrophy further indicate region-specific structural vulnerability rather than uniform involvement of related brain regions. Importantly, the weaker genetic coupling of atherosclerosis, despite its elevated burden in AD and VD, suggests that vascular pathology may be shaped more strongly by aging, inflammation, and environmental risk factors than by disorder-specific common genetic liability^79, 80^.

Many lines of evidence suggest that psychiatric disorders are not discretely unique disorders, but instead reflect overlapping, higher order symptom-dimensions^47, 81,82^. In line with this, we found that common neuropsychiatric symptoms, including hallucinations and delusions, were associated with distinct polygenic profiles, depending on the diagnostic context. In other words, a similar clinical outcome might have different underlying mechanisms, depending on the disease context. Although personality traits and sleep related genetic liability have previously been linked to mental health vulnerability and psychotic experiences^83–85^, their associations with hallucinations and or delusions in AD and PD have not, to the best of our knowledge, been described before. Moreover, the association of hallucinations and delusions with SCZ-PRS, but not to BP-PRS, is consistent with previous work showing that schizophrenia risk can relate to psychotic features^86,87^.

Several caveats warrant consideration. First, although this represents one of the largest neuropathologically characterized brain-bank cohorts to date, the sample size remains modest for genome-wide discovery, limiting power to detect novel loci and to perform direct locus-level comparisons with large public GWAS. Second, while we took measures to ensure that samples used for PRS calculation were not included in public GWAS, complete exclusion cannot be guaranteed. This overlap may have caused spurious p-value inflation within disorder-disorder comparisons (e.g. AD-group for AD-PRS). However, the cross-disorder associations, such as the significantly increased AD-PRS in MS, should be unaffected. Finally, the NBB, like other volunteer-based scientific cohorts, might exhibit selection bias. In addition, the cohort consists almost exclusively of donors of Dutch ancestry, which may limit the generalizability of our genetic findings to non-European ancestries.

Future efforts integrating multi-omic layers - including transcriptomic, methylation, and proteomic profiles - will be essential to bridge genetic variation with molecular and cellular pathology. Additionally, digital pathology will become critically important for automated characterization of a plethora of neuropathological endophenotypes. A major strength of this study is the integration of genetic data with rigorously defined post-mortem neuropathological diagnoses. By integrating genetic analyses with pathology, rather than with clinically defined diagnostics alone, we demonstrate that even a moderately sized cohort can refine established genetic associations and reveal previously unappreciated cross-disorder signals. Our results illustrate how neuropathologically informed cohorts can disentangle true pleiotropic risk from associations driven by clinical misclassification, thereby improving the interpretability of large-scale GWAS findings.

To facilitate further discovery, we made all polygenic risk scores available through our online resource (https://www.nnd.academy), together with detailed neuropathological diagnoses and symptom-level annotations. This resource enables researchers to investigate genetic risk across disorders, neuropathological endophenotypes, and neuropsychiatric symptoms groups. More broadly, we believe that it will be vitally important for other brain banks worldwide to harmonize and make multimodal data - genetic, clinical, and pathology and omics - accessible for large scale analysis and tissue selection^9,88^. Coordinated harmonization and open sharing of neuropathologically resolved genomic resources will empower the field to move beyond discrete symptom-based classification toward a biologically defined framework for understanding, diagnosing, and ultimately treating patients suffering from neurodegenerative, neuroinflammatory and psychiatric disorders. Together, the analyses and resources presented here may help guide the field toward an integrated, neuropathology-informed framework and support the development of more personalized approaches to diagnosis and treatment.

## Data Availability

All data produced in the present study are available upon reasonable request to the authors

https://nnd.academy/

## Acknowledgements

We would like to acknowledge Stephan Tap for annotating the neuropathology data under the supervision of Minke Groot. We are grateful to Johannes Schlachetzski for his careful reading of the manuscript and for providing extensive and constructive feedback. We would like to acknowledge Jörg Hamann and Erik Boddeke for their role in the NND executive board. Large language models were used solely for language editing, including grammar correction and refinement of individual sentences. No conceptual, analytical, or interpretative changes were made using AI. All authors reviewed and approved the final manuscript and take full responsibility for its content. We thank the investigators of the published GWASs from Schwartzentruber et al. (2021), Nalls et al. (2019), Chia et al. (2021, 2024), Ferrari et al. (2014), Wightman et al. (2023), van Rheenen et al. (2021), the International Multiple Sclerosis Genetics Consortium (IMSGC, 2019), Howard et al. (2019), Mullins et al. (2021), Trubetskoy et al. (2022), Shade et al. (2024), Schoeler et al. (2023), Savage et al. (2018), Bralten et al. (2021), and Hu et al. (2025) for making summary statistics available. These data were essential for the analyses performed in this study. Fig. 1a and Extended Data Fig. 1 were created with BioRender.com. Last but not least, we want to express our deepest gratitude toward the individuals who donated for the NBB and their family members.

## Author contributions

Conceptualization: I.R.H.

Methodology: N.J.M., S.K., I.R.H., E.H., H.J.W., K.L.K.

Formal analysis: N.J.M., S.K., I.R.H., E.H.

Investigation: N.J.M., S.K., I.R.H., A.R., E.H., E.D., A.M.G.

Resources: I.H., D.W., M.G., A.R., H.S., L.D.K., J.v.S., S.V., N.F., A.J.R.

Funding acquisition: I.R.H., I.H., B.J.L.E., A.J.R.

Supervision: I.R.H., B.J.L.E.

Writing – original draft: N.J.M., I.R.H., S.K., E.H.

Writing – review & editing: all authors.

## Methods

### The Netherlands Brain Bank

This study used data from donors of the NBB, with sample sizes varying by data modality: genetic data were available for 2,553 donors, clinical data for 2,339 donors, and neuropathological data for 2,667 donors (Fig. 1a; Supplementary Table 1). In short, the NBB collects brain tissue from individuals with neurological or psychiatric disorders, and controls donors, to facilitate brain research worldwide and improve the understanding of the human brain. All NBB procedures are in full compliance with Dutch and European law. The donors provided informed consent for the use of their tissue and their data for biomedical research purposes. The forms and procedures of the NBB were approved by the Free University Medical Center-Medical Ethics Committee (VUmc METC, Amsterdam, the Netherlands).

### Neuropathological examinations and neuropathological diagnosis groups

After each brain autopsy, neuropathologists from Amsterdam UMC pathology department performed extensive macroscopic and microscopic neuropathological examinations for the NBB. The neuropathologists assign a final diagnosis, which we referred to as ‘neuropathological diagnosis’ (Extended Data Table 1). For further information on all neuropathological diagnosis, we refer the reader to our previous publication^10^. Throughout this manuscript, diagnostic group labels refer to donors without a recorded secondary diagnosis, consistent with our previous work^10^. For example, donors with both AD and MS are classified as AD-MS, whereas AD refers exclusively to donors with a single AD diagnosis. The formal ND ontology underlying classification was previously published on Bioportal https://bioportal.bioontology.org/ontologies/NND_ND. For example, the AD group excludes donors with comorbid AD and PD diagnoses; donors with mixed AD-PD diagnoses are assigned to the AD-PD group. For the smaller FTD group, we included only donors diagnosed with FTD Picks’s disease, FTD-TDP subtypes, FTD-tau, FTD-FUS, or other-FTD (donors with multiple FTD-TDP subtypes, or n=1 subtypes such as FTD ubiquitin), while excluding donors with MND or additional neuropathological diagnoses. The psychiatric group includes donors with a psychiatric diagnosis such as MDD, BP, or SCZ. For the larger FTLD-MND spectrum group, comprising both different frontotemporal lobar degeneration disorders and motor neuron disease, we included all donors with FTD and/or MND, including donors with PSP, CBD, AGD, and mixed FTD-MND pathology.

### Clinical disease trajectories

We previously obtained clinical disease trajectories as described in^10^. Briefly, after death, the NBB staff retrieved all medical records, which were summarized, and translated into English. Natural language processing techniques were used to convert NBB medical record summaries into clinical disease trajectories encompassing 84 neuropsychiatric signs and symptoms.

### Extracting neuropathological endophenotypes using LLMs

Neuropathology reports from the NBB were stored in semi-standardized documents. These reports were parsed at sentence level, followed by cleaning and standardization of brain region terminology to ensure consistency across records. A subset of 280 randomly selected donor reports was manually annotated to serve as a reference dataset. Most neuropathological endophenotypes are recorded on an ordinal scale ranging from normal to severely affected, while features that could only be assessed as present or absent were recorded as a binary (Extended Data Table 3).

Feature extraction was performed using a LLM-based (Llama 3.1 8B Instruct^54^) pipeline (Extended Data Fig. 1a). First, relevant sentences for each endophenotype were identified using a combination of regular expression matching and fuzzy string (rapidfuzz 3.14.3)^89^ search. These candidate sentences were then processed using a carefully designed prompt to extract endophenotype values. To improve robustness, each sentence was evaluated 10 times by the LLM, and a consensus score was computed across predictions. For endophenotypes with less than 90% agreement on the annotated dataset, a secondary LLM was used for validation. This model flagged predictions (yes/no) and provided either a justification (if flagged) or the anchor term used for extraction (if not flagged). Subsequently, 100 randomly selected cases from both flagged and non-flagged groups were manually reviewed. Endophenotypes that failed to achieve ≥90% accuracy after this step were excluded from further analysis. The final dataset was compiled into a structured table with rows representing NND donor IDs and columns representing extracted neuropathological endophenotypes. In this study, only macroscopic and conclusion-level features were retained for downstream analysis. For macroscopic features, missing mentions were handled using a curation rule agreed with NBB neuropathologists: if a macroscopic abnormality was not reported, it was considered absent. For example, donors without a reported hippocampus atrophy were classified as non-atrophic. See Extended Data Table 3 for an overview of these neuropathological endophenotypes, their meaning, and scoring system.

### SNP Genotyping and quality control

DNA from 2,802 donors was isolated from postmortem blood, cerebellar tissue and genotyped on the Illumina GSA array V3. Hybridization, quality control and imputation were performed by the HUman GEnomics Facility (HUGE-F) of the Genetic Laboratory Rotterdam. Briefly, variants with excess heterozygosity (Hardy-Weinberg Equilibrium filter of 1*10^-5^) were removed, as were samples and variants with low call rate (< 97.5%). Variants were called with zCall^90^. Samples with low call rate (< 99%) or a sex mismatch, where the registered sex did not match the genetically inferred sex, were removed as well. Shape-it^91^ and Minimac4^92^ were used for phasing and imputation to the HRC reference panel^93^, respectively. To control for population structure in this mostly Dutch ancestry cohort, donors with (partial) non-European ancestries (first 4 PCs outside mean ±4SD based on PLINK1 --genome IBS/IBD analysis^94^) were removed. To avoid confounding, donors were removed if they had a family relationship (KING^95^) with another donor.

Before genetic association testing, we removed variants with Hardy–Weinberg equilibrium exact test p-value < 1*10^-5^, and with imputation info score < 0.8. PLINK2 v2.00^96^ sessions were generated with both minor allele frequency (MAF) ≥ 0.01 and MAF ≥ 0.05, suitable for polygenic risk score and genome-wide association study analyses, respectively. See Sup. Fig. 1A for an overview of the number of samples and variants passing each quality-control step.

### PCA and population stratification visualization

To assess the population structure and generate covariates for downstream analyses, PCA was conducted. PCA was performed on genotype data filtered at MAF ≥ 0.01 to maximize SNP coverage. Linkage disequilibrium (LD) pruning was applied (PLINK2 --indep-pairwise, excluding high LD regions) using a sliding window approach (window size 500kb), with SNPs exceeding a pairwise r^2^ threshold of 0.2 being removed.

To visualize the population stratification profile, we analyzed principal components (PCs) in relation to each donor’s last known province of residence, using residence as an approximate proxy for geographic ancestry, while recognizing that this may not reflect place of birth for all donors. We first used a Kruskal–Wallis test to identify PCs that were differentially distributed across Dutch provinces. PCs showing significant differences (Benjamini–Hochberg false discovery rate (FDR)–corrected p-values < 0.05) were then selected as input for a subsequent k-means clustering analysis, with the final number of seven clusters (k) determined using the elbow method. Subsequently, the k-means clusters were visualized on a choropleth map of the Netherlands constructed using GeoPandas (v1.0.1). Provinces were colored according to the normalized donor count (number of donors per 100,000 inhabitants). Pie charts were placed within the provincial boundaries representing the distribution of clusters within that province.

### Preprocessing of public GWAS summary statistics

GWAS summary statistics for different brain disorders and traits of interest were obtained and preprocessed. Disorders of interest include AD^30^, PD^31^, DLB^32^, FTD^33^, NDEG^34^, ALS^35^, MS^36^, MSA^37^, MDD^38^, BP^39^, and SCZ^40^. Neuropathological endophenotypes of interest^25^ included Amyloid-β plaques, Braak NFT stage, The Consortium to Establish a Registry for Alzheimer’s Disease (CERAD), Arteriolosclerosis, Atherosclerosis, CAA, Limbic-predominant age-related TDP-43 encephalopathy neuropathologic change (LATE-NC), and Hippocampal sclerosis. Traits of interest include insomnia and risk-taking^59^, intelligence^60^, sociability^61^, and neuroticism^62^. For an overview of GWASs used, see Extended Data Table 2. Summary statistics were converted to GrCh37, sorted by chromosome and position, and harmonized. GWASs were selected based on cohort size, and the presence of data needed for PRS calculation, being rsID or genetic coordinates, effect allele frequency, effect size or log odds ratio, standard error, p-value, and sample size per SNP. When the sample size per SNP was unavailable, the total sample size was used. Effect allele frequencies were inferred from the SBayesRC LD reference if missing. Missing standard errors were estimated as the natural logarithm of the odds ratio divided by z, with z approximated as the sign of the natural logarithm of the odds ratio multiplied by the absolute value of the quantile function applied to half of the p-value.

### Genetic correlations and gene-set enrichment analysis

Genetic correlations between GWASs were calculated using LDSC^42, 43^ with default settings and visualized as a heatmap. Gene-set enrichment analysis was performed using MAGMA^97^ with default parameters. Significant Gene Ontology Biological Process and Cellular Component gene sets representing key biological mechanisms were selected, and FDR corrected p-values were calculated and shown in a heatmap.

### Calculating polygenic scores

PRS were computed using SBayesRC^41^, which integrates GWAS summary statistics, LD structure, and functional annotations to estimate individual genetic risk in a target cohort. LD reference data were obtained from the UK Biobank European ancestry panel, and baseline model 2.2 was used for functional annotation^98^. To ensure compatibility with SBayesRC, the MAF>0.01 filtered imputed target genotype data underwent preprocessing, including SNP identifier harmonization with LD reference files. SNPs absent from the reference panel were excluded. PRS for each donor was calculated by using PLINK2 with the scoring file created with SBayesRC. Scores were centered to standardize the dosage values to zero. When comparing different PRS distributions across populations, scores were normalized using z-score normalization. We accounted for *APOE* effects in the AD PRS by first removing the region around the *APOE* gene (chr19: 44409039-46412650) and then adding the *APOE* effect, modeled based on ε2 + ε4 risk haplotype dose, as previously published^99^. For neuropathological endophenotype PRS^25^, variants within the *APOE* locus were excluded from score construction to reduce the influence of this single high-impact region. *APOE* ε2/ε4 haplotype dosage was then retained as a separate genetic score to assess *APOE*-specific associations.

### Analysis of polygenic scores

To investigate which neuropathological diagnoses groups were enriched for specific disease associated PRSs, we compared the distribution of PRS z-score of each group to the control group with 10.000 permutation tests, followed by an FDR correction. The results were visualized using a dot plot, where dot color indicated the median z-score difference in PRS between the disorder group and the control group. An asterisk was used to indicate significant differences (FDR corrected p-value < 0.05), with the size of each dot corresponding to the FDR-corrected p-value.

### FTLD-MND spectrum mutation status and polygenic risk scores

To investigate the influence of familial mutations on the FTLD-MND spectrum, we obtained familial mutation status for a subset of donors^52^. We compiled and standardized this information at the gene level of known familial mutations in the *MAPT, GRN, C9orf72* gene. We subsequently tested whether these different mutation groups differed in the FTD, AD, ALS and PD – PRS using an FDR corrected permutation test.

### Association between polygenic scores and neuropsychiatric symptoms

To investigate whether polygenic risk contributes to symptom-level heterogeneity within neuropathologically defined disorders, we tested the association between polygenic scores for disorders and personality traits with neuropsychiatric symptom observation (yes/no). Analyses were performed in four diagnostic groups (AD, PD, Control, psychiatric). We selected nine neuropsychiatric symptoms with minimal prevalence of 10 observations and nine polygenic scores, including PRS for the disorders AD, MDD, BP, SCZ, and polygenic scores for the personality traits risk-taking, intelligence, neuroticism, sociability, and insomnia. For each diagnostic group, we fitted logistic regression models to assess the association between standardized PRS and binary symptom presence, adjusting for the first three principal components as previously published^86^. To account for multiple testing and non-independence between traits, empirical significance thresholds were determined using 10,000 phenotype-label permutations per PRS–symptom pair, controlling the false positive rate (α = 0.005).

### GWAS refinement through GWAS on NND samples

To evaluate whether loci identified in the Schwartzentruber AD meta-GWAS-by-proxy could be replicated in the AD donors of our neuropathologically defined cohort, or were potentially driven by other neurodegenerative disorders such as PD or FTD, we conducted three case–control GWASs for AD (N-controls=276, N-cases=553), FTD (N-controls=276, N-cases=193), and PD (N-controls=283, N-cases=238) using Plink2. GWASs were performed with the logistic-Firth hybrid regression model to account for modest sample sizes. Age, sex, and the first three genotype PCs were included as covariates, with covariate variance standardization applied. AD summary statistics from Schwartzentruber were segmented into loci as defined in the original publication and when needed extended to a minimum region of 1Mb around the lead SNP. Loci were excluded if the lead SNP p-value was larger than 1e-10. NND summary statistics were segmented in the same way and harmonized with the SNPs reported in the public summary statistics. Colocalization analyses^53^ were performed to evaluate whether signals in the ADSchwartzentruber and NND GWAS were consistent with a shared causal variant. Loci with a posterior probability for a shared causal variant (PP4) greater than 0.8 were considered colocalized. PP4 values between pairs of GWAS were visualized as a heatmap. As a secondary validation, for the locus of interest we also extracted the original summary statistics and controlled the FDR using the Benjamini–Hochberg procedure as previously published^25^ where variants with an adjusted Q value ≤ 0.05 were considered significant. Candidate loci with potential for refinement were visualized using stacked regional association plots (GENI plots, version 0.1.2).

### Analysis of neuropathological endophenotypes

To quantify disease-specific pathological burden, we first calculated the average burden of each extracted neuropathological endophenotype within each disease category and in controls. For ordinal features, burden was defined as the mean feature score; for binary features, burden was defined as the proportion of donors with the feature present. Differences in burden were then calculated relative to the control group and normalized by the maximum possible value of the corresponding feature to allow comparison across scoring scales. Statistical differences between each disease group and controls were assessed using a two-sided Mann-Whitney U test for ordinal features and Fisher’s exact test for binary features. P-values were adjusted across comparisons using the Benjamini-Hochberg FDR procedure.

We then tested whether variation in these neuropathological endophenotypes was associated with genetic risk by calculating associations with PRSs. Because the extracted neuropathological endophenotypes were encoded as ordinal or binary variables, Spearman’s rank correlation was used to measure monotonic associations without assuming normal distributions. Each neuropathological endophenotype - PRS pair was analyzed independently using donors with complete data for that specific comparison. Statistical significance was assessed using a two-sided permutation test (10,000 permutations). The observed correlation was compared against true distribution to estimate p-values. To account for multiple testing across all neuropathological endophenotypes and PRS combinations, p-values were adjusted using the Benjamini–Hochberg FDR procedure. Adjusted p-values < 0.05 were considered statistically significant.

Second, we calculated PRSs using summary statistics from GWASs of multiple neuropathological endophenotypes^25^. Only summary statistics with at least one genome-wide significant signal were used.

---

**Ext. 1.**
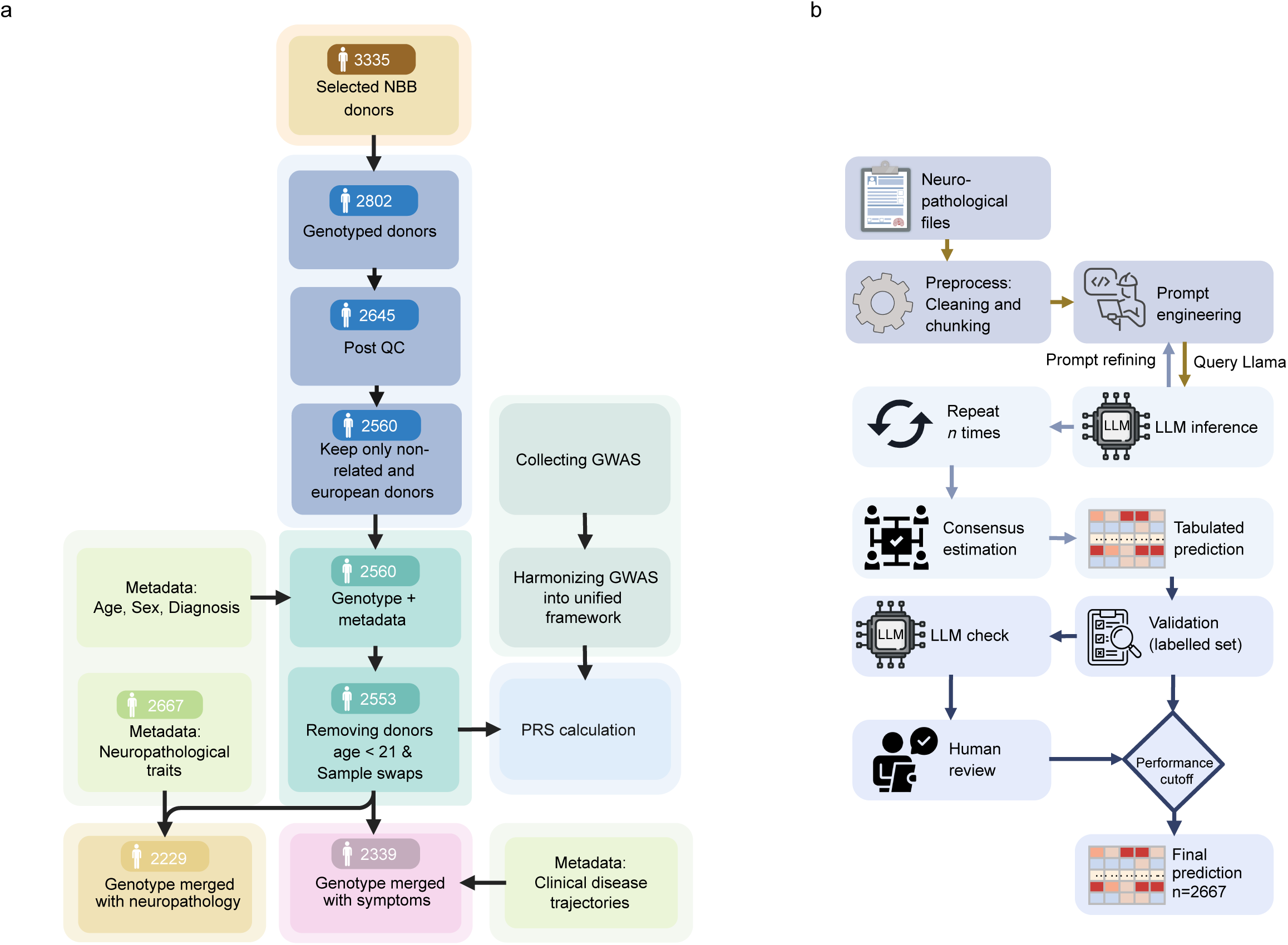
a, Overview of genotyping workflow. Genotype data undergoes QC (e.g. removing samples with low call rate or excess heterozygosity) and removal of non-european and related donors, sample swaps, and donors under age 21. PRS are calculated based on GWAS summary statistics. Metadata is added. b, LLM-based pipeline for extracting neuropathological endophenotypes. Relevant sentences were identified using regex and fuzzy matching, followed by LLM-based feature extraction with consensus across multiple runs. Low-confidence predictions were validated using a secondary LLM and manual review. Features not reaching ≥90% accuracy were excluded.

**Ext. 2.**
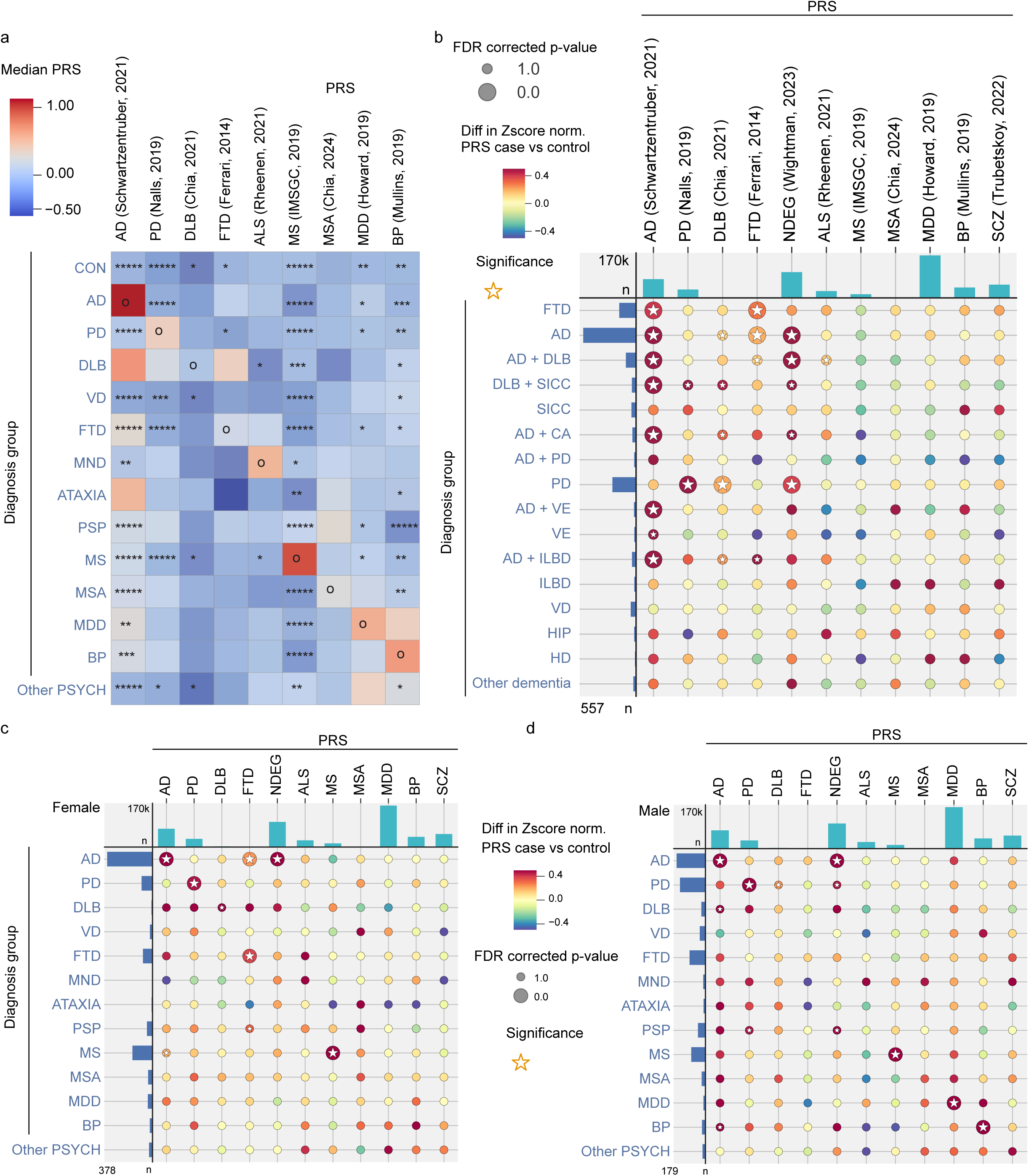
a, Heatmap of median standardized PRS across diagnostic groups (rows for 9 disorders (columns). Color indicates median PRS. Circles (o) denote the diagnostic group selected as the reference for each PRS; this group is compared against all other groups within the same column using permutation testing with FDR correction. Asterisks indicate significance (*P < 0.05, **P < 0.005, ***P < 0.0005, ****P < 0.00005, *****P < 0.000005). b, PRS for 11 different disorders (columns) for 16 rare and mixed dementias (rows). Color indicates the direction and magnitude of the difference in standardized PRS (case vs control). Dot size reflects statistical significance, and stars denote FDR-corrected permutation test P < 0.05. Column bar plot indicates GWAS sample size (n cases). Row bar plots indicate NND sample size. c-d, dotplot as in Fig. 2 d, but stratified for female (c) and male (d) donors respectively. Legend for c and d is shared.

**Ext. 3.**
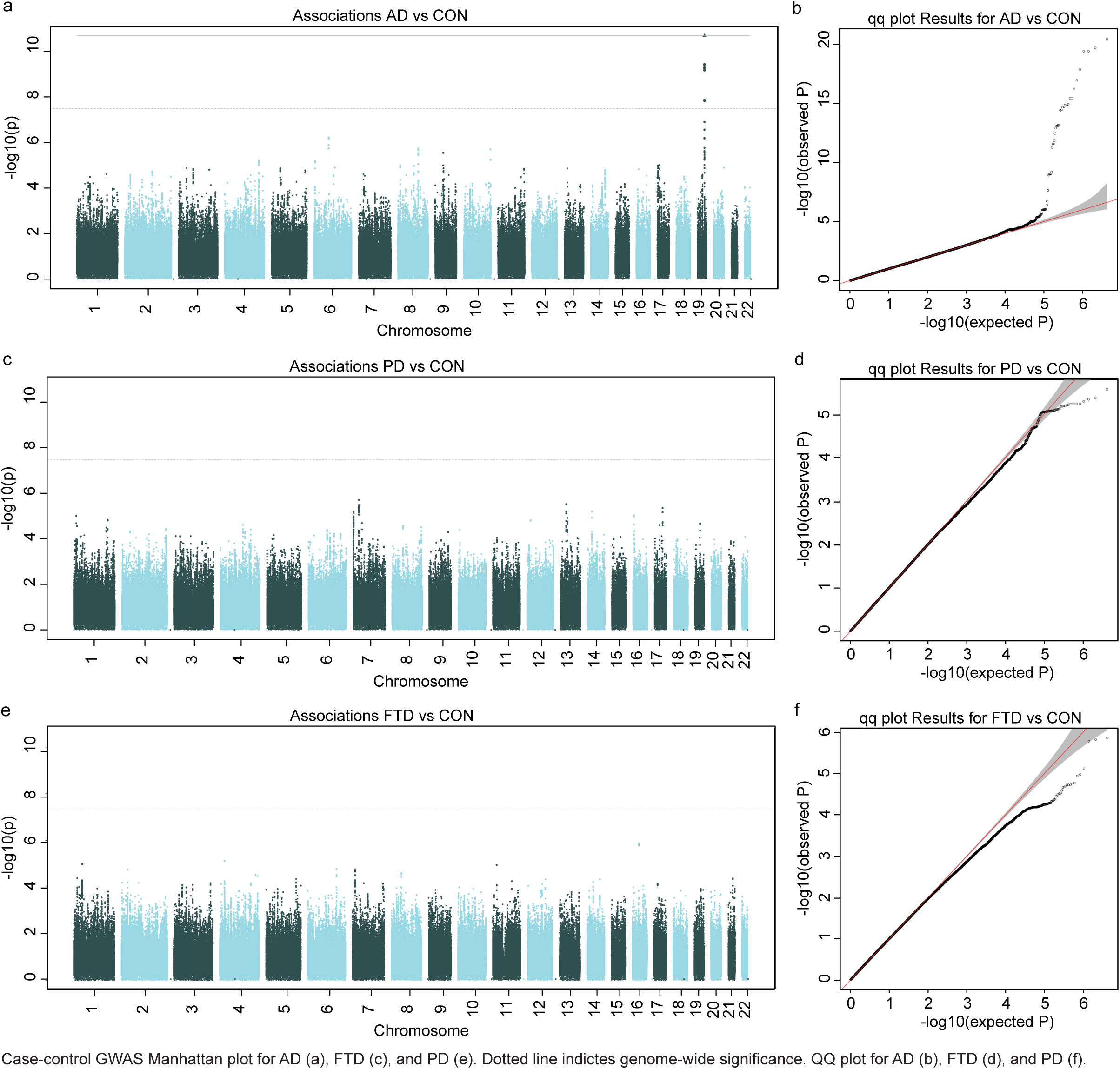
Case-control GWAS Manhattan plot for AD (a), FTD (c), and PD (e). Dotted line indictes genome-wide significance. QQ plot for AD (b), FTD (d), and PD (f).

**Ext. 4.**
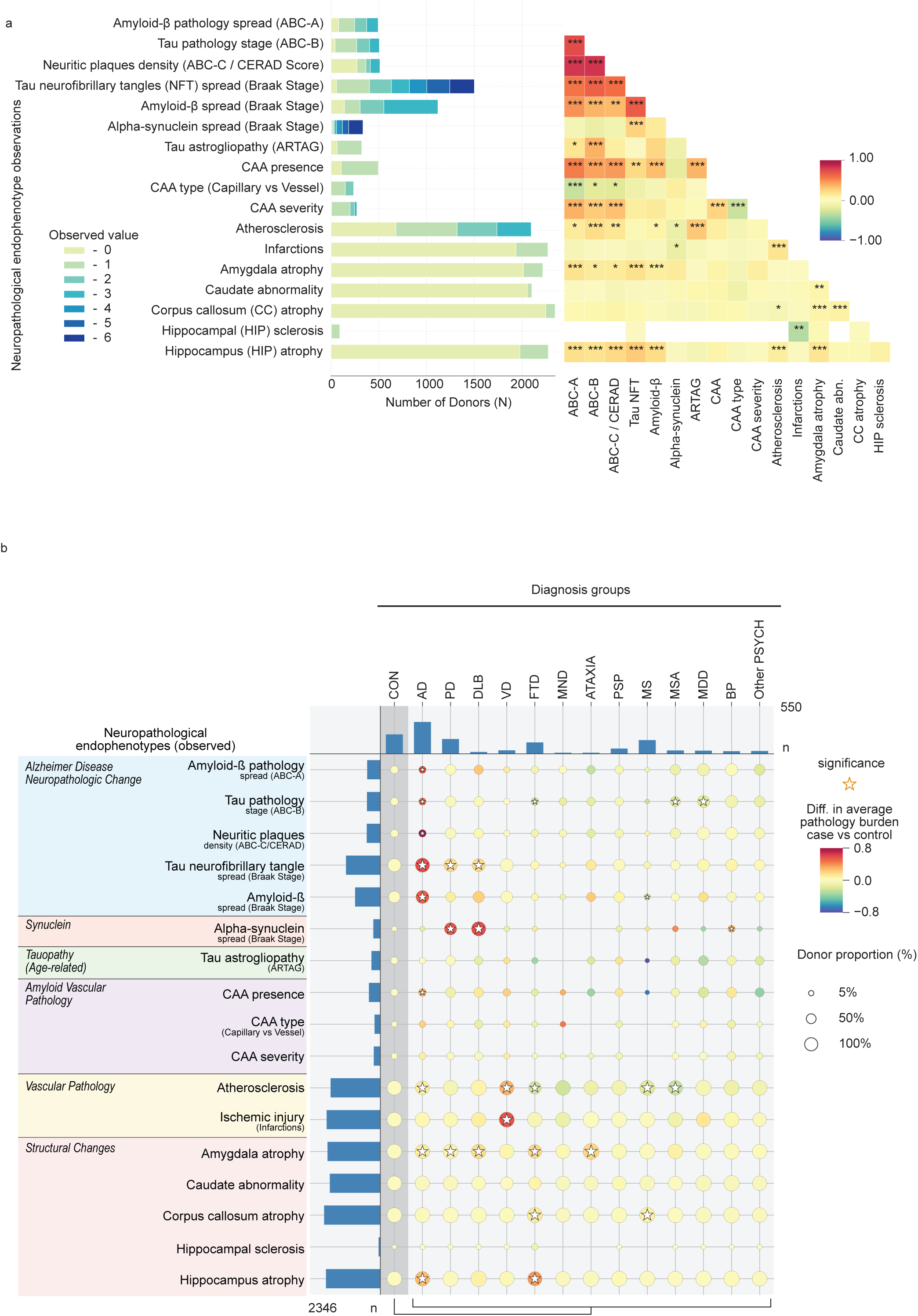
Quantitative landscape of neuropathological endophenotypes and polygenic risk. a, Stacked bar plot showing the distribution of 17 extracted neuropathological endophenotypes across the cohort, with colors indicating observed scores. Adjacent heatmap shows pairwise Spearman correlations between neuropathological endophenotypes. Color indicates the direction and magnitude of correlation, and stars denote FDR-corrected significance levels. No. of stars denote significance levels: * P<0.05, ** P<0.01, *** P<0.001. b, Dot plot showing the difference in average burden of each neuropathological endophenotype across 13 neuropathological diagnostic groups relative to controls. Burden was calculated as the mean score for ordinal features and as the proportion of positive cases for binary features. Color intensity indicates the magnitude and direction of the difference from controls. Differences were tested using two-sided Mann-Whitney U tests for ordinal features and Fisher’s exact tests for binary features, followed by Benjamini–Hochberg FDR correction. Dot size represents the percentage of donors with an observation, and stars indicate significant differences from controls (P adj <0.05).

**Extended Data Table 1.** | Overview of neuropathological diagnoses, sample size, and additional information.

| Neuropathological diagnosis | n | Additional information |
| --- | --- | --- |
| Control Brain (CON) | 281 | Donors that do not meet the criteria for any brain disorder |
| Alzheimer's disease (AD) | 557 | ICD10 G30 |
| Parkinson's disease (PD) | 249 | ICD10 G20 |
| Multiple sclerosis (MS) | 255 | ICD10 G35 |
| Multiple system atrophy (MSA) | 59 | MESH D019578 |
| Vascular dementia (VD) | 56 | ICD10 F01 |
| Dementia with Lewy bodies (DLB) | 33 | MESH D020961 |
| Frontotemporal Dementia (FTD) | 177 | ICD10 G31.0 Includes FTD-TDP, FTD-FUS, Pick's disease, and others |
| Motor neuron disease (MND) | 18 | ICD10 G12.2 Includes ALS and other motor neuron diseases |
| FTD-TDP + MND | 19 | Donors with both an FTD-TDP and MND diagnosis |
| Cerebellar Ataxia (ATAXIA) | 18 | ICD10 G11 |
| Argyrophilic grain disease (AGD) | 11 | SNOMEDCT 230281007 |
| Corticobasal degeneration (CBD) | 16 | ICD10 G31.85 |
| Progressive supranuclear palsy (PSP) | 92 | MESH D013494 |
| Bipolar disorder (BP) | 43 | ICD10 F31 |
| Major depressive disorder (MDD) | 51 | ICD F33 |
| Other psychiatric disorders (Other PSYCH) | 46 | Includes schizophrenia (ICD10 F20), PTSD (ICD10 F43.1), and others |
| Vascular encephalopathy (VE) | 18 | Donors with neuropathological vascular lesions (infarctions/atherosclerosis/hemorrhages) |
| Huntington's disease (HD) | 12 | ICD10 G10 |
| Hippocampal sclerosis (HIP) | 20 | ICD10 G93.81 |
| Incidental Lewy Body Disease (ILBD) | 19 | Incidental Lewy body disease is the presence of Lewy bodies or Lewy neurites without clinical findings of parkinsonism or dementia |
| Senile Involutive Cortical Changes (SICC) | 48 | Progress of amyloid plaques and neurofibrillary tangles, but not sufficient to meet the criteria for Alzheimer's disease |
| Other Dementia (DEM other) | 29 | Includes donors with multiple dementias |
| Brain Cancer | 14 | Includes neoplasms of the brain, both primary (benign/malignant) and secondary (metastatic) forms. |
| Tauopathy | 10 | MESH D024801 |
| Vascular pathology | 37 | Includes ischemic changes, cerebrovascular accident, and others |
| Other Neuropath | 10 | Includes congophilic angiopathy, Creutzfeldt-Jakob disease, and others |
| AD + CA | 38 | Donors with both an AD and CA diagnosis |
| AD + DLB | 107 | Donors with both an AD and DLB diagnosis |
| AD + ILBD | 43 | Donors with both an AD and ILBD diagnosis |
| AD + PD | 20 | Donors with both an AD and PD diagnosis |
| AD + VE | 22 | Donors with both an AD and VE diagnosis |
| DLB + SICC | 44 | Donors with both an DLB and SICC diagnosis |

**Extended Data Table 2.** | Overview of GWAS used in this manuscript, including accompanying publication and sample size.

| GWAS | Publication | PMID | n |
| --- | --- | --- | --- |
| Alzheimer's disease (AD) | Schwartzentruber et al. (2021) | 33589840 | 472,868 |
| Parkinson's disease (PD) | Nalls et al. (2019) | 31701892 | 482,730 |
| Dementia with Lewy bodies (DLB) | Chia et al. (2021) | 33589841 | 6,618 |
| Frontotemporal dementia (FTD) | Ferrari et al. (2014) | 24943344 | 6,462 |
| Neurodegenerative disorder (NDEG) | Wightman et al. (2023) | 37045620 | 1,025,492 |
| Amyotrophic lateral sclerosis (ALS) | van Rheenen et al. (2021) | 34873335 | 138,086 |
| Multiple sclerosis (MS) | IMSGC (2019) | 31604244 | 41,505 |
| Multiple system atrophy (MSA) | Chia et al. (2024) | 38701790 | 8,016 |
| Major depressive disorder (MDD) | Howard et al. (2019) | 30718901 | 500,199 |
| Bipolar disorder (BP) | Mullins et al (2021) | 34002096 | 413,466v |
| Schizophrenia (SCZ) | Trubetskoy et al (2022) | 35396580 | 130,644 |
| Amyloid-beta plaques | Shade et al. (2024) | 39379761 | 6,363 |
| Braak NFT Stage | Shade et al. (2024) | 39379761 | 7,776 |
| CERAD | Shade et al. (2024) | 39379761 | 7,786 |
| Arteriolosclerosis | Shade et al. (2024) | 39379761 | 6,668 |
| Atherosclerosis | Shade et al. (2024) | 39379761 | 7,340 |
| CAA | Shade et al. (2024) | 39379761 | 7,381 |
| LATE-NC | Shade et al. (2024) | 39379761 | 3,112 |
| Hippocampal sclerosis | Shade et al. (2024) | 39379761 | 7,164 |
| Risk taking | Schoeler et al. (2023) | 37106081 | 274,537 |
| Intelligence | Savage et al. (2018) | 29942086 | 269,867 |
| Insomnia | Schoeler et al. (2023) | 37106081 | 283,595 |
| Sociability score | Bralten et al. (2021) | 34054130 | 342,461 |
| Neuroticism | Hu et al. (2025) | 39901012 | 279,645 |

**Extended Data Table 3.** | Overview of the Neuropathological endophenotypes used in this manuscript, including explanation, scale, and sample size.

| Name | Classification | Explanation | Type | Scale | n |
| --- | --- | --- | --- | --- | --- |
| Amyloid-β pathology spread (ABC-A) | Alzheimer Disease Neuropathologic Change | Derived from Thal phase | Ordinal | None = 0, cortex = 1, striatum/thalamus = 2, brainstem/cerebellum = 3 | 493 |
| Tau pathology stage (ABC-B) |  | Based on Braak stage for NFT | Ordinal | No NFTs = 0, Stage 1–2 = 1, Stage 3–4 = 2, Stage 5–6 = 3 | 506 |
| Neuritic plaques density (ABC-C/CERAD Score) |  | Density of neuritic plaques based on CERAD scoring | Ordinal | None = 0, Mild/Slight = 1, Moderate = 2, Severe = 3 | 512 |
| Tau neurofibrillary tangles spread (Braak Stage) |  | Distribution of tau neurofibrillary tangle pathology across brain regions | Ordinal | None = 0, Transentorhinal = 1(0.17), Entorhinal = 2, Fusiform/lingual neocortex = 3, Association neocortex = 4, Peristriate neocortex = 5, Primary/secondary neocortex = 6 | 1501 |
| Amyloid-β spread (Braak Stage) | | Distribution of amyloid-beta deposition across brain regions | Ordinal | None = 0, Basal isocortex = A, Association isocortex = B, Primary sensory/motor cortex = C | 1119 |
| Alpha-synuclein spread (Braak Stage) | Synucleinopathy | Distribution of α-synuclein/Lewy body pathology across brain regions | Ordinal | None = 0, Medulla = 1, Pons = 2, Midbrain/basal forebrain/hippocampus CA2 = 3, Basal forebrain/hippocampus/transentorhinal cortex = 4, Cingulate gyrus/temporal cortex = 5, Frontal/parietal cortex = 6 | 334 |
| Tau astroglial pathology (ARTAG) | Tauopathy (Age-related) | Astrocytic tau accumulation associated with aging | Binary | Absent = 0, Present = 1 | 322 |
| CAA presence | Amyloid Vascular Pathology | Amyloid deposition in cerebral vessel walls | Binary | Absent = 0, Present = 1 | 494 |
| CAA type (Capillary vs Vessel) |  | Type 1 involves amyloid deposition in capillaries; Type 2 affects larger vessels. | Categorical | Capillary = 1, Vessel = 2 | 237 |
| CAA severity |  | Degree of vascular amyloid burden | Ordinal | None = 0, Mild/Slight = 1, Moderate = 2, Severe = 3 | 270 |
| Atherosclerosis | Vascular Pathology | Thickening and narrowing of cerebral arteries due to plaque build up | Ordinal | None = 0, Mild/Slight = 1, Moderate = 2, Severe = 3 | 2095 |
| Infarctions |  | Areas of brain tissue loss due to lack of blood flow | Binary | Absent = 0, Present = 1 | 2270 |
| Amygdala atrophy | Structural Changes | Reduction in tissue volume of amygdala | Binary | Absent = 0, Present = 1 | 2217 |
| Caudate abnormality |  | Flattening of the caudate | Binary | Absent = 0, Present = 1 | 2102 |
| Corpus callosum atrophy |  | Reduction in tissue volume of corpus callosum | Binary | Absent = 0, Present = 1 | 2346 |
| Hippocampal sclerosis |  | Severe neuronal loss and gliosis in hippocampus | Binary | Absent = 0, Present = 1 | 92 |
| Hippocampus atrophy |  | Reduction in tissue volume of hippocampus | Binary | Absent = 0, Present = 1 | 2274 |

## References

1. You, J., Guo, Y. et al. Clinical trajectories preceding incident dementia up to 15 years before diagnosis: a large prospective cohort study. Mol Psychiatry 29, 3097–3105 (2024).

2. Lian, J. et al. Subtyping Alzheimer’s disease and Parkinson’s disease using longitudinal electronic health records. Nat Aging 6, 612–625 (2026).

3. Giebel, C. et al. “A systematic review on the evidence of misdiagnosis in dementia and its impact on accessing dementia care.” Int J Geriatr Psychiatry, 39, e6158 (2024).

4. Brownlee, W. J. & Solomon, A. J. “Misdiagnosis of multiple sclerosis: Time for action.” Mult. Scler. 27(6), 805–806 (2021).

5. Mukwikwi, E. R., et al. “Prevalence and Features of Misdiagnosis of Primary Psychiatric Disorders Among bvFTD Patients.” J Neuropsychiatry Clin Neurosci, 37(4), 364–370 (2025).

6. Kentrou, V. et al. “Perceived misdiagnosis of psychiatric conditions in autistic adults.” EClinicalMedicine 71, 102586 (2024).

7. Young, A.L., Oxtoby, N.P. et al. Data-driven modelling of neurodegenerative disease progression: thinking outside the black box. Nat. Rev. Neurosci. 25, 111–130 (2024).

8. Shin, D., Lee, S. et al. Biomarker-integrated prognostic stagings for Alzheimer’s Disease. Nat Commun 17, 2235 (2026).

9. Murray, M.E. et al. “Accelerating biomedical discoveries in brain health through transformative neuropathology of aging and neurodegeneration.” Neuron 113(22), 3703–3721 (2025).

10. Mekkes, N.J. et al. “Identification of clinical disease trajectories in neurodegenerative disorders with natural language processing.” Nat Med 30, 1143–1153 (2024).

11. Robinson, J.L. et al. “Pathological combinations in neurodegenerative disease are heterogeneous and disease-associated.” Brain 146(6), 2557–2569 (2023).

12. Jellinger, K.A. “Recent update on the heterogeneity of the Alzheimer’s disease spectrum.” J Neural Transm 129(1), 1–24 (2022).

13. Zhang, M. et al. “The correlation between neuropathology levels and cognitive performance in centenarians.” Alzheimers Dement. 19(11), 5036–5047 (2023).

14. Smith, J.E. Most dementia patients have multiple brain diseases. How should they be treated? Science news: Brain & Behavior (2026). Available at: https://www.science.org/content/article/most-dementia-patients-have-multiple-brain-diseases-how-should-they-be-treated (accessed 27 May 2026)

15. Van Rumund, A. et al. “α-Synuclein real-time quaking-induced conversion in the cerebrospinal fluid of uncertain cases of parkinsonism.” Ann Neurol 85(5), 777–781 (2019).

16. Holtman I.R., Glass C.K., Nott A. “Interpretation of Neurodegenerative GWAS Risk Alleles in Microglia and their Interplay with Other Cell Types.” Adv Neurobiol 37, 531–544 (2024).

17. Uffelmann, E. et al. “Genome-wide association studies.” Nat Rev Methods Primers 1, 59 (2021)

18. Keogh, M.J., et al. “Genetic compendium of 1511 human brains available through the UK medical research council brain banks network resource.” Genome Res 27(1), 65–173 (2017).

19. Smeland, O.B., et al. “The shared genetic risk architecture of neurological and psychiatric disorders: a genome-wide analysis.” medRxiv (2023).

20. Stolp Andersen, M., et al. “Dissecting the limited genetic overlap of Parkinson’s and Alzheimer’s disease.” Annals of clinical and translational neurology 9(8), 1289–1295 (2022).

21. Reynolds, R.H. et al. “Local genetic correlations exist among neurodegenerative and neuropsychiatric diseases.” npj Parkinson’s Disease 9(1), 70 (2023).

22. Wainberg, M., Andrews, S.J., & Tripathy, S.J. “Shared genetic risk loci between Alzheimer’s disease and related dementias, Parkinson’s disease, and amyotrophic lateral sclerosis.” Alzheimer’s Research & Therapy 15(1), 113 (2023).

23. Koretsky, M.J. et al. “Genetic risk factor clustering within and across neurodegenerative diseases.” Brain 146(11), 4486–4494 (2023).

24. Wu, Y. et al. “Pervasive biases in proxy genome-wide association studies based on parental history of Alzheimer’s disease.” Nat. Genet. 56(12), 2696–2703 (2024).

25. Shade, L. M. P. et al. “GWAS of multiple neuropathology endophenotypes identifies new risk loci and provides insights into the genetic risk of dementia.” Nat. Genet. 56(11), 2407–2421 (2024).

26. Nott, A. et al. “Brain cell type–specific enhancer–promoter interactome maps and disease-risk association.” Science 366(6469), 1134–1139 (2019).

27. Hardy, J. & Escott-Price, V. “The genetics of neurodegenerative diseases is the genetics of age-related damage clearance failure.” Mol. Psychiatry 30(6), 2748–2753 (2025).

28. International Multiple Sclerosis Genetics Consortium, et al. “Multiple sclerosis genomic map implicates peripheral immune cells and microglia in susceptibility.” Science 365(6460), eaav7188 (2019).

29. Byrne, R. P. et al. “Dutch population structure across space, time and GWAS design.” Nat. Commun. 11(1), 4556 (2020).

30. Schwartzentruber, J. et al. “Genome-wide meta-analysis, fine-mapping and integrative prioritization implicate new Alzheimer’s disease risk genes.” Nat. Genet. 53(3), 392–402 (2021).

31. Nalls, M. A. et al. “Identification of novel risk loci, causal insights, and heritable risk for Parkinson’s disease: a meta-analysis of genome-wide association studies.” Lancet Neurol. 18(12), 1091–1102 (2019).

32. Chia, R. et al. “Genome sequencing analysis identifies new loci associated with Lewy body dementia and provides insights into its genetic architecture.” Nat. Genet. 53(3), 294–303 (2021).

33. Ferrari, R. et al. “Frontotemporal dementia and its subtypes: a genome-wide association study.” Lancet Neurol. 13(7), 686–699 (2014).

34. Wightman, D. P. et al. “The genetic overlap between Alzheimer’s disease, amyotrophic lateral sclerosis, Lewy body dementia, and Parkinson’s disease.” Neurobiol. Aging 127, 99–112 (2023).

35. Van Rheenen, W. et al. “Common and rare variant association analyses in amyotrophic lateral sclerosis identify 15 risk loci with distinct genetic architectures and neuron-specific biology.” Nat. Genet. 53(12), 1636–1648 (2021).

36. International Multiple Sclerosis Genetics Consortium, et al. “Multiple sclerosis genomic map implicates peripheral immune cells and microglia in susceptibility.” Science 365(6460), eaav7188 (2019).

37. Chia, R. et al. “Genome sequence analyses identify novel risk loci for multiple system atrophy.” Neuron 112(13), 2142–2156 (2024).

38. Howard, D. M. et al. “Genome-wide meta-analysis of depression identifies 102 independent variants and highlights the importance of the prefrontal brain regions.” Nat. Neurosci. 22(3), 343–352 (2019).

39. Mullins, N. et al. “Genome-wide association study of more than 40,000 bipolar disorder cases provides new insights into the underlying biology.” Nat. Genet. 53(6), 817–829 (2021).

40. Trubetskoy, V. et al. “Mapping genomic loci implicates genes and synaptic biology in schizophrenia.” Nature 604(7906), 502–508 (2022).

41. Zheng, Z. et al. “Leveraging functional genomic annotations and genome coverage to improve polygenic prediction of complex traits within and between ancestries.” Nat. Genet. 56(5), 767–777 (2024).

42. Bulik-Sullivan, B. K. et al. “LD Score regression distinguishes confounding from polygenicity in genome-wide association studies.” Nat. Genet. 47(3), 291–295 (2015).

43. Bulik-Sullivan, B. et al. “An atlas of genetic correlations across human diseases and traits.” Nat. Genet. 47(11), 1236–1241 (2015).

44. Miedema, A. et al. “Brain macrophages acquire distinct transcriptomes in multiple sclerosis lesions and normal appearing white matter.” Acta Neuropathol. Commun. 10(1), 8 (2022).

45. LaCroix, M. S. et al. “Tau seeding in cases of multiple sclerosis.” Acta Neuropathol. Commun. 10(1), 146 (2022).

46. Nowacki, P., Koziarska, D. & Masztalewicz, M. “Microglia and astroglia proliferation within the normal appearing white matter in histologically active and inactive multiple sclerosis.” Folia Neuropathol. 57(3), 249–257 (2019).

47. Grotzinger, A. D. et al. “Genomic structural equation modelling provides insights into the multivariate genetic architecture of complex traits.” *Nat*. Hum. Behav. 3(5), 513–525 (2019).

48. Thomas, J. T. et al. “Sex-stratified genome-wide association meta-analysis of major depressive disorder.” Nat. Commun. 16(1), 7960 (2025).

49. Pottier, C. et al. “Genetics of FTLD: overview and what else we can expect from genetic studies.” J. Neurochem. 138, 32–53 (2016).

50. Saez-Atienzar, S. et al. “Mechanism-free repurposing of drugs for C9orf72-related ALS/FTD using large-scale genomic data.” Cell Genom. 4(11), 100679 (2024).

51. Mars, N. et al. “The role of polygenic risk and susceptibility genes in breast cancer over the course of life.” Nat. Commun. 11(1), 6383 (2020).

52. Mol, M. O. et al. “Underlying genetic variation in familial frontotemporal dementia: sequencing of 198 patients.” Neurobiol. Aging 97, 148.e9–148.e16 (2021).

53. Giambartolomei, C. et al. “Bayesian test for colocalisation between pairs of genetic association studies using summary statistics.” PLoS Genet. 10(5), e1004383 (2014).

54. Grattafiori, Aaron, et al. “The llama 3 herd of models.” arXiv preprint arXiv:2407.21783 (2024).

55. Richard, Edo, Melina GHE den Brok, and Willem A. van Gool. “Bayes analysis supports null hypothesis of anti-amyloid beta therapy in Alzheimer’s disease.” Alzheimer’s & Dementia 17(6), 1051–1055 (2021).

56. Richard, Edo, et al. “The Alzheimer myth and biomarker research in dementia.” Journal of Alzheimer’s Disease 31(3), S203–S209 (2012).

57. van der Molen, Lennart H., et al. “Changing definitions of disease: Transformations in the diagnostic criteria for Alzheimer’s disease.” Alzheimer’s & Dementia 21(4), e70133 (2025).

58. Maristany, A. J., et al. “Psychiatric manifestations of neurological diseases: a narrative review.” Cureus 16(7), e64152 (2024).

59. Schoeler, T. et al. “Participation bias in the UK Biobank distorts genetic associations and downstream analyses.” *Nat*. Hum. Behav. 7(7), 1216–1227 (2023).

60. Savage, J. E. et al. “Genome-wide association meta-analysis in 269,867 individuals identifies new genetic and functional links to intelligence.” Nat. Genet. 50(7), 912–919 (2018).

61. Bralten, J. et al. “Genetic underpinnings of sociability in the general population.” Neuropsychopharmacology 46(9), 1627–1634 (2021).

62. Hu, S. et al. “Fine-scale population structure and widespread conservation of genetic effect sizes between human groups across traits.” Nat. Genet. 57(2), 379–389 (2025).

63. DeMichele-Sweet, M. A. A. et al. “Genome-wide association identifies the first risk loci for psychosis in Alzheimer disease.” Mol. Psychiatry 26(10), 5797–5811 (2021).

64. Creese, B. et al. “Examining the association between genetic liability for schizophrenia and psychotic symptoms in Alzheimer’s disease.” Transl. Psychiatry 9(1), 273 (2019).

65. Escott-Price, V. & Hardy, J. “Genome-wide association studies for Alzheimer’s disease: bigger is not always better.” Brain Commun. 4(3), fcac125 (2022).

66. Emeršič, A. et al. “Biomarkers of tau phosphorylation state are associated with the clinical course of multiple sclerosis.” Mult. Scler. Relat. Disord. 90, 105801 (2024).

67. Anderson, J. M., et al. “Abnormally phosphorylated tau is associated with neuronal and axonal loss in experimental autoimmune encephalomyelitis and multiple sclerosis.” Brain 131(7), 1736–1748 (2008).

68. Anderson, Jane Marian, et al. “Abnormal tau phosphorylation in primary progressive multiple sclerosis.” Acta neuropathologica 119(5), 591–600 (2010).

69. LaCroix, Michael S., et al. “Tau seeding in cases of multiple sclerosis.” Acta Neuropathologica Communications 10(1), 146 (2022).

70. Fominykh, V. et al. “Shared genetic loci between Alzheimer’s disease and multiple sclerosis: Crossroads between neurodegeneration and immune system.” Neurobiol. Dis. 183, 106174 (2023).

71. Fuchs TA, et al. Cognitive progression independent of relapse in multiple sclerosis. Mult Scler. 30(11-12):1468–1478 (2024).

72. Van Der Ende, E. L. et al. “Unravelling the clinical spectrum and the role of repeat length in C9ORF72 repeat expansions.” J. Neurol. Neurosurg. Psychiatry 92(5), 502–509 (2021).

73. Murray, M. E. et al. “Clinical and neuropathologic heterogeneity of c9FTD/ALS associated with hexanucleotide repeat expansion in C9ORF72.” Acta Neuropathol. 122(6), 673–690 (2011).

74. Devenney, E. et al. “Clinical heterogeneity of the C9orf72 genetic mutation in frontotemporal dementia.” Neurocase 21(4), 535–541 (2015).

75. Kingdom, R. et al. “Genetic modifiers of rare variants in monogenic developmental disorder loci.” Nat. Genet. 56(5), 861–868 (2024).

76. Moss, D. J. H. et al. “Identification of genetic variants associated with Huntington’s disease progression: a genome-wide association study.” Lancet Neurol. 16(9), 701–711 (2017).

77. Tai, M.-C. et al. “Polygenic risk score for breast cancer risk prediction in Asian BRCA1 and BRCA2 pathogenic variants carriers.” NPJ Breast Cancer 11(1), 105 (2025).

78. Owens, H. A. et al. “Alzheimer’s disease-associated P460L variant of EphA1 dysregulates receptor activity and blood-brain barrier function.” Alzheimers Dement. 20(3), 2016–2033 (2024).

79. Lathe, Richard, Alexandra Sapronova, and Yuri Kotelevtsev. “Atherosclerosis and Alzheimer-diseases with a common cause? Inflammation, oxysterols, vasculature.” BMC geriatrics 14(1), 36 (2014).

80. Wang, Jukun, Jing Yao, and Zhe Wang. “Identification of shared mechanisms between Alzheimer’s disease and atherosclerosis by integrated bioinformatics analysis.” European Journal of Medical Research 30(1), 408 (2025).

81. Kotov, R. et al. “The Hierarchical Taxonomy of Psychopathology (HiTOP): A quantitative nosology based on consensus of evidence.” Annu. Rev. Clin. Psychol. 17(1), 83–108 (2021).

82. Grotzinger, A. D. et al. “Mapping the genetic landscape across 14 psychiatric disorders.” Nature 649(8096), 406–415 (2026).

83. Holt-Lunstad, J. “Social connection as a critical factor for mental and physical health: evidence, trends, challenges, and future implications.” World Psychiatry 23(3), 312–332 (2024).

84. Jeronimus, B. F. et al. “Neuroticism’s prospective association with mental disorders halves after adjustment for baseline symptoms and psychiatric history, but the adjusted association hardly decays with time: a meta-analysis on 59 longitudinal/prospective studies with 443,313 participants.” Psychol. Med. 46(14), 2883–2906 (2016).

85. Longenecker, J. M., Krueger, R. F. & Sponheim, S. R. “Personality traits across the psychosis spectrum: a hierarchical taxonomy of psychopathology conceptualization of clinical symptomatology.” Pers. Ment. Health 14(1), 88–105 (2020).

86. Coombes, B. J. et al. “Dissecting clinical heterogeneity of bipolar disorder using multiple polygenic risk scores.” Transl. Psychiatry 10(1), 314 (2020).

87. Richards, A. L. et al. “Genetic liabilities differentiating bipolar disorder, schizophrenia, and major depressive disorder, and phenotypic heterogeneity in bipolar disorder.” JAMA Psychiatry 79(10), 1032–1039 (2022).

88. Samarasekera, N. et al. “Brain banking for neurological disorders.” Lancet Neurol. 12(11), 1096–1105 (2013).

## Method references

89. Bachmann, Max, et al. “rapidfuzz/RapidFuzz: Release 3.14.3.” Zenodo (2025).

90. Goldstein, J. I. et al. “zCall: a rare variant caller for array-based genotyping: genetics and population analysis.” Bioinformatics 28(19), 2543–2545 (2012).

91. Delaneau, J., Marchini, J. & Zagury, J.-F. “A linear complexity phasing method for thousands of genomes.” Nat. Methods 9(2), 179–181 (2012).

92. Das, S. et al. “Next-generation genotype imputation service and methods.” Nat. Genet. 48(10), 1284–1287 (2016).

93. McCarthy, S. et al. “A reference panel of 64,976 haplotypes for genotype imputation.” Nat. Genet. 48, 1279–1283 (2016).

94. Purcell, S. et al. “PLINK: a tool set for whole-genome association and population-based linkage analyses.” Am. J. Hum. Genet. 81(3), 559–575 (2007).

95. Manichaikul, A. et al. “Robust relationship inference in genome-wide association studies.” Bioinformatics 26(22), 2867–2873 (2010).

96. Chang, C. C. et al. “Second-generation PLINK: rising to the challenge of larger and richer datasets.” Gigascience 4, 7 (2015).

97. De Leeuw, C. A. et al. “MAGMA: generalized gene-set analysis of GWAS data.” PLoS Comput. Biol. 11(4), e1004219 (2015).

98. Márquez-Luna, C. et al. “Incorporating functional priors improves polygenic prediction accuracy in UK Biobank and 23andMe data sets.” Nat. Commun. 12(1), 6052 (2021).

99. Leonenko, G. et al. “Identifying individuals with high risk of Alzheimer’s disease using polygenic risk scores.” Nat. Commun. 12(1), 4506 (2021).

